# Minimum changes in sleep, physical activity, and nutrition associated with clinically important reductions in all-cause mortality risk: a prospective cohort study

**DOI:** 10.1101/2023.11.19.23298747

**Authors:** Emmanuel Stamatakis, Nicholas A. Koemel, Raaj K. Biswas, Matthew N. Ahmadi, Margaret Allman-Farinelli, Stewart G. Trost, Elif I. Eroglu, Borja del Pozo Cruz, Yu Sun Bin, Svetlana Postnova, Stephen Simpson, Mitch Duncan, Dorothea Dumuid, Luigi Fontana, Helen Brown, Carol Maher, Peter A. Cistulli

**Affiliations:** Mackenzie Wearables Research Hub, Charles Perkins Centre, The University of Sydney, Sydney, New South Wales, Australia; School of Health Sciences, Faculty of Medicine and Health, The University of Sydney, Sydney, New South Wales, Australia; Charles Perkins Centre, The University of Sydney, Sydney, New South Wales, Australia; Nutrition and Dietetics, School of Nursing, Faculty of Medicine and Health, The University of Sydney, Sydney, New South Wales, Australia; School of Human Movement and Nutrition Sciences, The University of Queensland, and Children’s Health Queensland, Brisbane, Queensland, Australia; German Institute of Human Nutrition Potsdam-Rehbruecke, Department of Molecular Epidemiology, Nuthetal, Germany; Faculty of Education, Department of Physical Education, University of Cádiz, Cádiz, Spain; Biomedical Research and Innovation Institute of Cádiz (INiBICA) Research Unit, University of Cádiz, Cádiz, Spain; Department of Sports Science and Clinical Biomechanics, University of Southern Denmark, Odense, Denmark; School of Physics, Faculty of Science, The University of Sydney, Sydney, New South Wales, Australia; School of Life and Environmental Sciences, Faculty of Science, The University of Sydney, Sydney, New South Wales, Australia; School of Medicine and Public Health; College of Health, Medicine, and Wellbeing, The University of Newcastle, University Drive, Callaghan, New South Wales, Australia; Centre for Active Living and Learning, University of Newcastle, Callaghan, New South Wales, Australia; Alliance for Research in Exercise, Nutrition and Activity, Allied Health and Human Performance, University of South Australia, Adelaide, South Australia, Australia; Department of Endocrinology, Royal Prince Alfred Hospital, Sydney, New South Wales, Australia; School of Exercise and Nutrition Sciences, Faculty of Health, Deakin University, Geelong, Victoria, Australia; Department of Respiratory and Sleep Medicine, Royal North Shore Hospital, Sydney, New South Wales, Australia

**Keywords:** Sleep, nutrition, physical activity, mortality, cohort studies

## Abstract

**Background:** Sleep, physical activity, and nutrition (SPAN) are crucial modifiable factors for health, yet most research has examined them independently rather than exploring their combined and incremental impact on disease risk and mortality.

**Objective:** To determine the collective associations of SPAN exposures and establish clinically relevant targets for reducing all-cause mortality risk.

**Methods:** This study included 59,078 UK Biobank participants with valid wearable tracker and nutrition data (Median age [IQR]: 64.0 [7.8] years; 45.4% male). Sleep duration (hours/day) and moderate to vigorous physical activity duration (MVPA; mins/day) were calculated using a machine learning based wearable data schema. A 10-item diet quality score (DQS) assessed the consumption of vegetables, fruits, fish, dairy, whole grains, and vegetable oils, as well as lower intakes of refined grains, processed meats, unprocessed red meats, and sugar-sweetened beverages using a food frequency questionnaire. The DQS assigned values from 0-10 for each component, totalling 100 points, with higher values indicating higher diet quality. Associations with all-cause mortality were explored using Cox proportional hazard models with combinations of SPAN exposure tertiles.

**Results:** During the median 8.1-year follow-up period, 2,458 deaths occurred. MVPA exhibited the strongest overall effect on mortality risk, followed by sleep (with a U-shaped relationship), and diet quality. Compared to the referent group of combined SPAN exposure (lowest tertiles for all three behaviours), the optimal SPAN combination involving moderate sleep duration (7.2-8.0 hours/day), high MVPA (42-103 mins/day), and high DQS (57.5-72.5) was associated with a hazard ratio (HR) of 0.45 (95% CI: 0.37, 0.53). Relative to the 5th percentile of sleep (5.5 hours/day), physical activity (7.3 mins/day), and nutrition (36.9 DQS), a minimum increase of 15 mins/day of sleep, 1.6 min/day MVPA, and 5 DQS points was associated with a 10% reduction in all-cause mortality risk (HR: 0.90; 95% CI: 0.88, 0.93). Additionally, compared to the referent group, an additional 75 mins/day of sleep, 12.5 min/day MVPA, and 25 DQS points was associated with a 50% reduction in all-cause mortality risk (HR: 0.50; 95%CI: 0.44, 0.58).

**Conclusion:** These findings underscore the importance of combined incremental lifestyle modifications in reducing the risk of all-cause mortality.

## INTRODUCTION

Adequate sleep, physical activity, and nutrition (SPAN) are vital for health and well-being, and each has established links with chronic disease and mortality. Both under and over-sleeping impair metabolic and brain health through mechanisms such as insulin resistance, inflammation, and disruption of appetite hormones [1–3]. Physical inactivity undeniably contributes to the promotion of multiple chronic diseases [4], and adhering to current guidelines is linked with a 29% lower risk for all-cause mortality [5]. Excessive calorie intake and poor nutrition play a key role in the pathogenesis of some of the most common non-communicable diseases and in the biology of aging itself [6, 7]. Aspects of diet quality such as the intake of ultra-processed food and sugar-sweetened beverages, refined grains, processed meat, along with low intake of fruit, vegetables, whole grains, beans and fish have some of the strongest links between disease risk and metabolic health [8]. Sleep, physical activity, and nutrition are behaviourally interlinked, often clustering together to form broader lifestyle patterns [9, 10]. For instance, sleep deprivation can lead to lower physical activity due to subjective fatigue and reduced time to exhaustion, while exercise uptake may lead to improvements in sleep [11–14]. Poor sleep upregulates signalling for hunger hormones and downregulates satiety hormones, directly influencing food intake and food choices [15]. Additionally, low-quality diets negatively impact sleep by disrupting the neurotransmitters that regulate normal sleep-wake cycles [16].

Despite this high degree of interdependency, the three SPAN behaviours have been studied and promoted mostly in silos. Moreover, research that examines the optimal approaches to tackling these key chronic disease risk factors is scarce. A small number of studies have explored joint associations of pairs of these behaviours with all-cause, cardiovascular disease, and cancer mortality, for example diet and physical activity [17, 18], or sleep and physical activity [19, 20]. A small study of just over 1000 National Health and Nutrition Examination Survey 2005-2006 participants revealed the effects of different combinations of the SPAN exposures with all-cause mortality *via* model-based clustering analysis [21]. This study revealed unique behavioural SPAN profiles were associated with mortality. Specifically, lowest risk for all-cause mortality was observed in individuals with average diet quality, high physical activity, low sedentary behaviour, and average sleep. An analysis of self-reported data from The 45 and Up Study also highlighted that the combined effect of over-sleeping, physical inactivity, excessive sedentary time, and poor dietary patterns are likely to be larger than the sum of risk associated with each individual behaviour [22].

This emerging body of evidence suggests interventions addressing multiple SPAN behaviours concurrently may have promising potential for reducing mortality risk. However, there is no information on the minimal amount of change required for each behaviour to observe a measurable combined synergistic difference in health outcomes. Sustainable SPAN behaviour change is challenging, particularly through the traditional intervention approaches that usually set targets for substantial changes in one of the three SPAN behaviours. No study has examined the minimum, and hence more likely behaviourally sustainable, improvements across all three SPAN behaviours required for measurable improvements in health outcomes.

The first aim of this study was to examine the combined associations of SPAN exposures with the risk of all-cause mortality in a large UK cohort using device-measured sleep and physical activity and a comprehensive diet quality score for nutrition. Our second aim was to identify the minimum change in sleep, physical activity and diet associated with clinically meaningful reductions in all-cause mortality risk.

## METHODS

### Study Population

We used data from the UK Biobank cohort study which recruited 502,629 adults ages 40-69 from 2006 to 2010 [23]. Participants completed touchscreen questionnaires for sociodemographic, lifestyle characteristics, and health status. All participants completed informed consent and the ethical approval was completed by the UK National Health service (NHS) and National Research Ethics Service for the UK (No. 11/NW/0382).

Dietary data was derived using a self-report twenty-nine item food-frequency questionnaire, collected at recruitment (2006-2009) to determine the frequency of commonly consumed foods and food groups over the past 12-month period [24]. During 2013 to 2015, a subgroup of 103,684 participants were mailed and wore wrist-worn accelerometers (Axivity AX3, York, UK) on their dominant wrist for 7-days. As previously described, participants were only included in the present study if they had wrist-worn accelerometry data collected with sufficient wear time of at least three days (>16 hours/day) with one of the days being a weekend day [25–28]. Participants were also excluded from the primary analysis if no sleep data was recorded, accelerometer was poorly calibrated, or a faulty accelerometer was distributed. We also excluded participants who reported they were unable to walk or with incomplete covariate information (**Supplemental Figure 1**).

### Outcome Ascertainment

Participants were followed through November 30^th^, 2022, with all-cause mortality information including follow-up duration and mortality events retrieved via the data linkage program from the National Health Service Central Register and National Records of Scotland. To avoid potential reverse causation, we excluded all participants with a mortality event in the first year of follow-up [25, 27].

### Sleep, Physical Activity, and Nutrition

Sleep information was derived from accelerometry data and defined as the average daily duration of sleep (hours/day). The duration of sleep was calculated using a validated algorithm based on relative changes in wrist tilt angle between successive 5 second windows [29, 30]. For each interval of 5 seconds, the average of the estimated wrist tilt angle was calculated and served as an input for the algorithm. Sleep periods were identified by a tilt angle change of less than 5 degrees for 5 minutes. Physical activity was explored using accelerometer-derived daily minutes of moderate to vigorous physical activity (MVPA). All accelerometers were calibrated and initialized to 100 Hz and participants were instructed to wear the device on their dominant wrist for 7 consecutive days. The intensity of physical activity was defined using a previously validated two-stage machine learning approach which classifies activities into light, moderate, or vigorous physical activity in 10 second windows [25, 26, 31, 32].

From the recorded dietary information, a diet quality score (DQS) was extracted which emphasizes higher intake of vegetables, fruits, fish, dairy, whole grains, and vegetable oils and lower intake of refined grains, processed meats, unprocessed red meats, and sugar-sweetened beverages [33]. This score rates the intake of each category on a scale from 0 (unhealthiest) to 10 (healthiest) for a total of 100 points, where higher values equate to a higher diet quality.

### Statistical Analyses

We estimated the associations between SPAN lifestyle characteristics and all-cause mortality using Cox proportional hazard regression models. To minimize the influence of sparse data and outliers, the values below the 2.5 percentile and above the 97.5 percentile for all SPAN exposures were truncated in this analysis. We first plotted dose-response plots for each SPAN exposure to explore the minimum dose of SPAN exposures needed to have a clinically meaningful reduction in all-cause mortality risk. As per previous work, [34, 35] we defined clinically meaningful changes in all-cause mortality risk as a 10% reduction in risk or the nearest predicted integer. To offer context on associations of each SPAN behaviour with all-cause mortality risk in individuals of differing behavioural status, we show dose-response plots for using both the 5th percentile (considered ‘unhealthy’) and median (considered ‘healthy’) as reference points while adjusting for the SPAN behaviours not examined (**Supplemental Figure 2**). To create a more granular table for clinically meaningful interpretations, we plotted a heatmap correlogram that uses the 5^th^ percentile of all SPAN exposures as reference while providing a corresponding hazard ratio for incremental changes of each exposure (i.e., 10^th^, 30^th^, 50^th^, 70^th^, and 90^th^ percentile rounded to nearest 5^th^ integer).

Participants then were grouped by SPAN exposure tertiles (i.e., low, moderate, and high) which equated to a joint exposure of 27 separate groups for all three SPAN behaviours. The specific ranges for each exposure included sleep duration as 4.8-7.2 hours/day (low), 7.2-8.0 hours/day (medium), and 8.0-9.4 hours/day (high); MVPA measurements as 5-23 mins/day (low), 23-42 mins/day (medium), and 42-103 mins/day (high); and diet quality using the DQS as 32.5-50.0 (low), 50.0-57.5 (medium), and 57.5-72.5 (high). In these models, we included each SPAN exposure as an independent term to explore combined effects. We first explored the associations of SPAN exposures with all-cause mortality by plotting each of the 27 categories as a forest plot with the lowest tertile of each exposure used as the reference point (**Figure 1**). Model estimates were adjusted for age, sex, ethnicity, smoking, education, Townsend deprivation index, alcohol, discretionary screen time (time spent watching TV or using the computer outside of work), light intensity physical activity, medication (blood pressure, insulin, and cholesterol), previous diagnosis of major CVD (defined as disease of the circulatory system, arteries, and lymph, excluding hypertension), previous diagnosis of cancer, and familial history of CVD and cancer. Further details regarding these covariates are provided in **Supplemental Table 1**. All models satisfied proportional hazards assumptions using Schoenfeld residuals.

**Figure 1:**
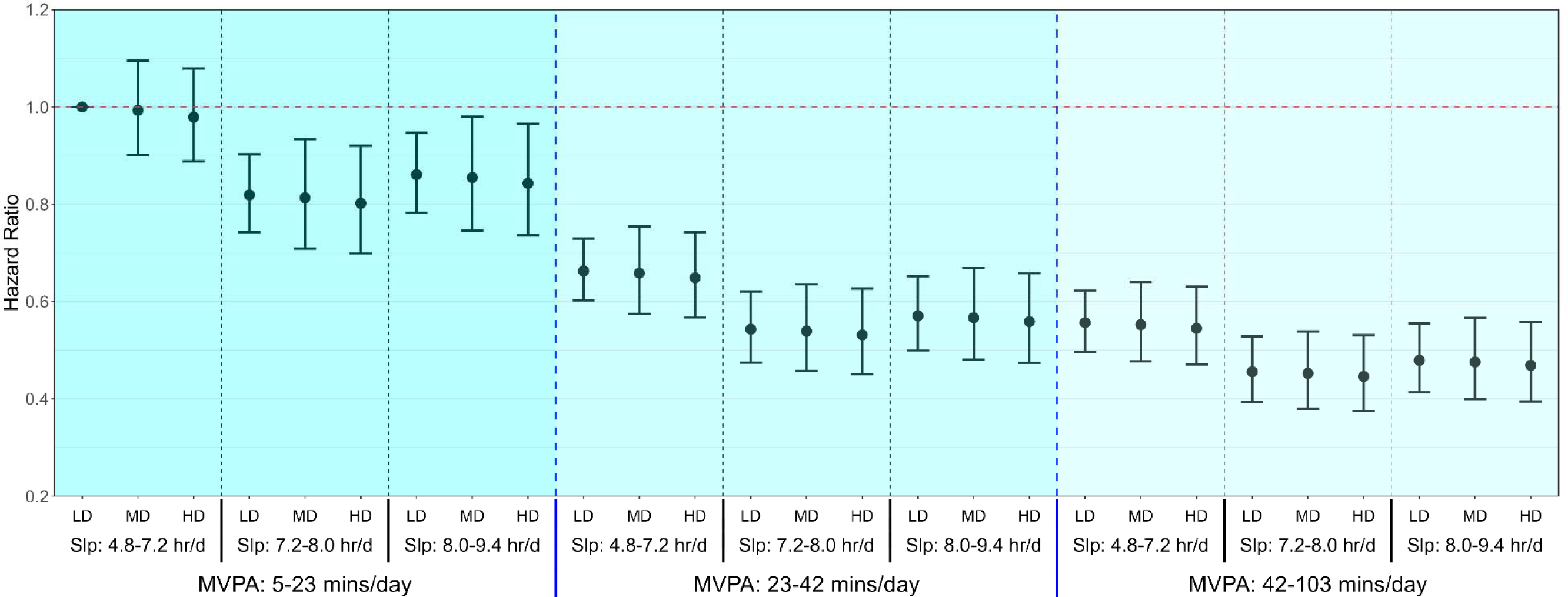
Association of sleep, MVPA, and diet quality with all-cause mortality Model is adjusted for age, sex, ethnicity, smoking, education, Townsend deprivation index, alcohol, discretionary screen time (time spent watching TV or using the computer outside of work), light intensity physical activity, medication (blood pressure, insulin, and cholesterol), previous diagnosis of major CVD (defined as disease of the circulatory system, arteries, and lymph, excluding hypertension), previous diagnosis of cancer, and familial history of CVD and cancer. Dashed blue lines separate tertiles MVPA and dashed black lines separate tertiles of sleep. Moderate-Vigorous Physical Activity (MVPA); Low Diet Quality (LD); Medium Diet Quality (MD); High Diet Quality (HD).

### Sensitivity Analyses

We excluded individuals who self-reported poor health, current smokers, those in the top 20th percentile of the frailty index, and those with an underweight BMI (<18.5). We also adjusted the model for BMI, which may be a mediator in the association between each SPAN behaviour and mortality.

We undertook all statistical analyses and visualisations using the survival, rms, ggplot2 packages of R (version 4.3.1). We followed the Strengthening the Reporting of Observational Studies in Epidemiology (STROBE) guidelines (**Supplemental Table 3**).

## RESULTS

### Sample

The final analytic sample included 59,078 participants (Median age [IQR]: 64.0 [7.8] years; 45.4% male) and 2,458 all-cause mortality events (**Table 1**). After excluding events in the first 12 months of follow-up, the average follow-up period was 8.1 [7.5-8.6] years. Median [IQR] sleep for each tertile was 6.52 [6.89, 8.26], 7.62 [7.41, 7.83], and 8.52 [8.26, 8.89] hours/day for low, medium, and high sleepers. For physical activity, medians were 14.31 [9.64, 18.52], 31.26 [26.76, 36.26], and 58.74 [49.09, 74.91] mins/day of MVPA for low, medium, and high tertiles. Median diet quality scores (DQS) were 45.00 [40.00, 47.50], 55.00 [52.50, 56.79], and 62.50 [60.00, 67.50] for low, medium, and high tertiles.

**Table 1:**
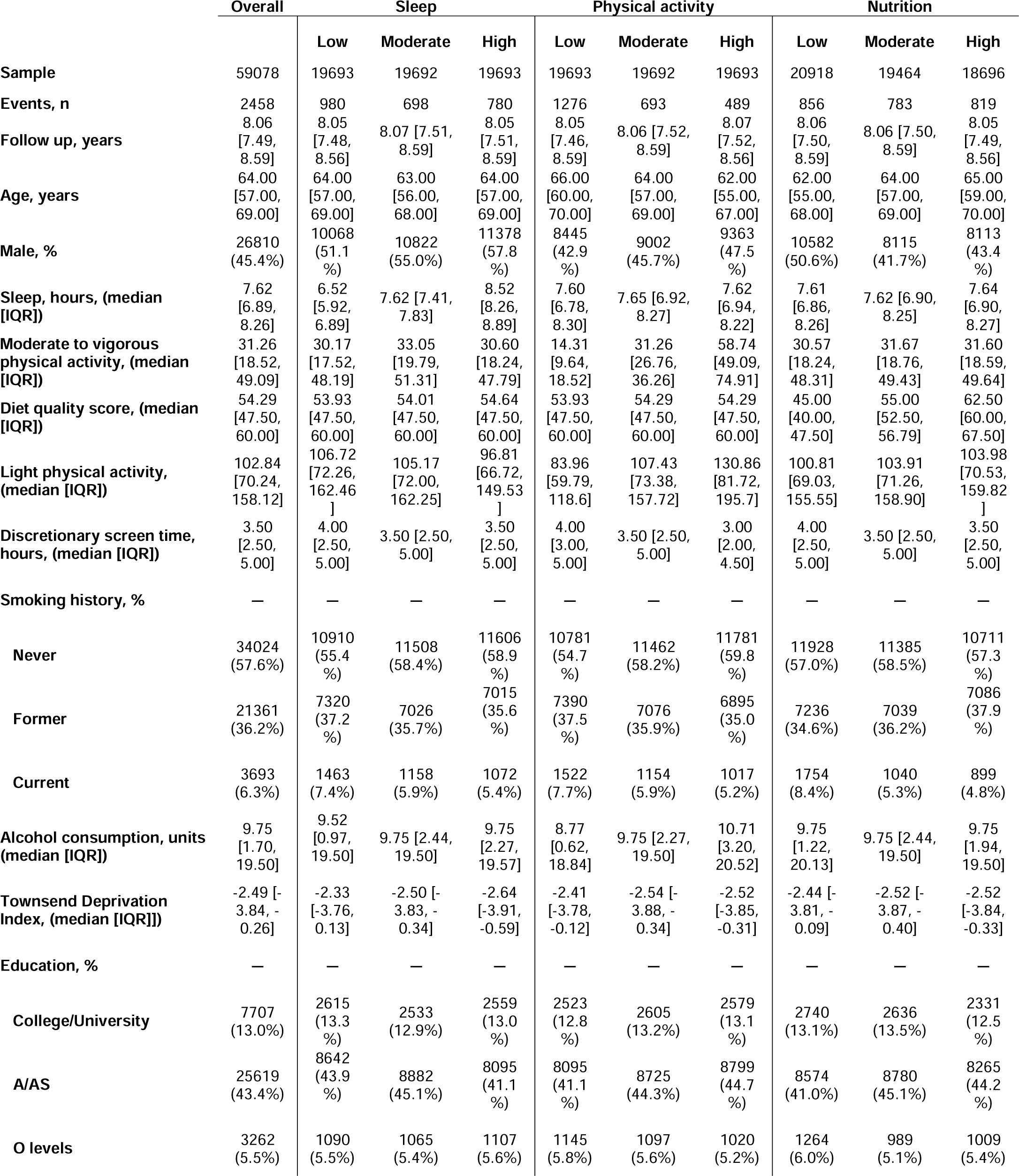
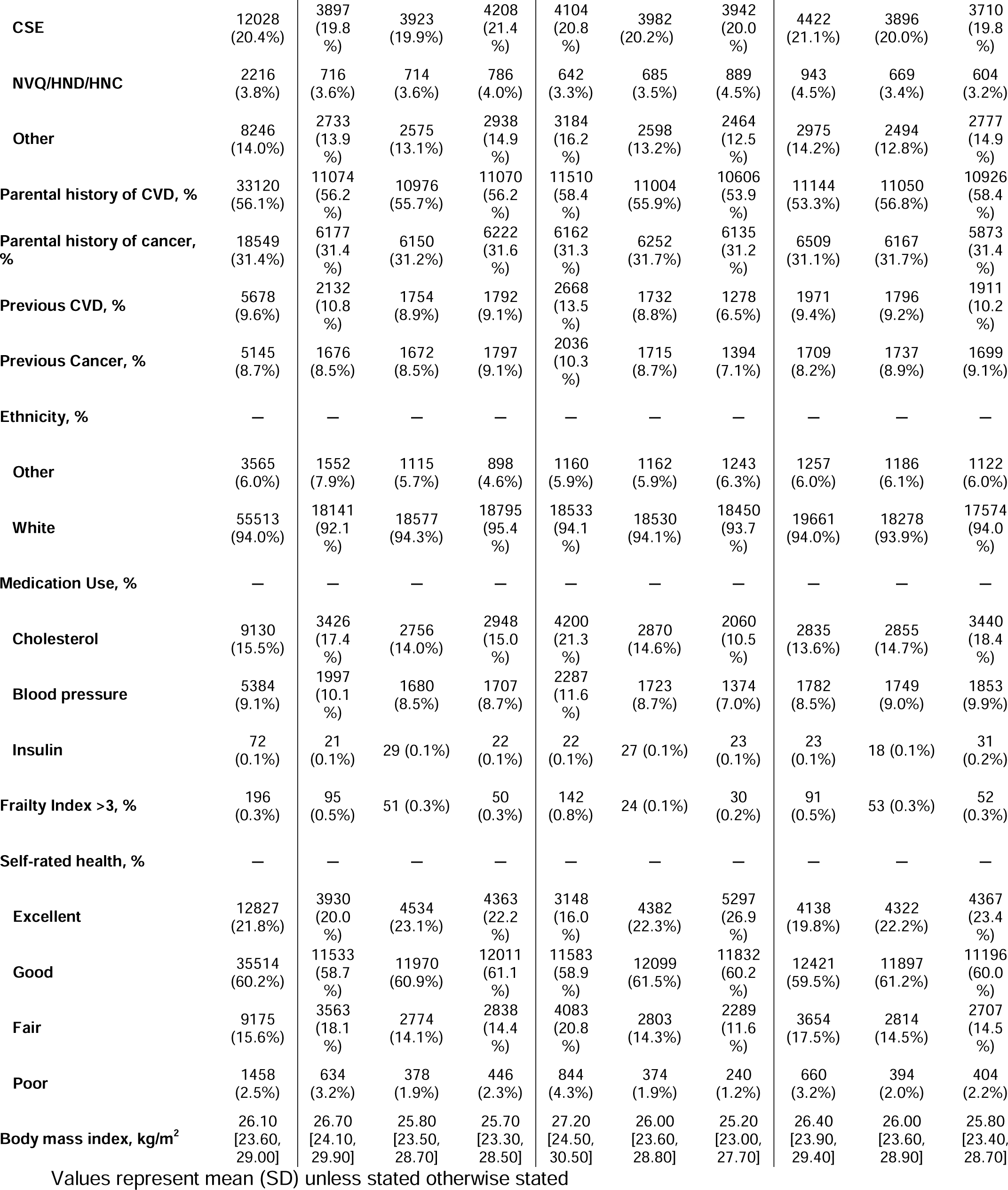
Participant characteristics.

|  | Overall | Sleep |  |  | Physical activity |  |  | Nutrition |  |  |
| --- | --- | --- | --- | --- | --- | --- | --- | --- | --- | --- |
|  |  | Low | Moderate | High | Low | Moderate | High | Low | Moderate | High |
| Sample | 59078 | 19693 | 19692 | 19693 | 19693 | 19692 | 19693 | 20918 | 19464 | 18696 |
| Events, n | 2458 | 980 | 698 | 780 | 1276 | 693 | 489 | 856 | 783 | 819 |
| Follow up, years | 8.06<br>[7.49, 8.59] | 8.05<br>[7.48, 8.56] | 8.07 [7.51, 8.59] | 8.05<br>[7.51, 8.59] | 8.05<br>[7.46, 8.59] | 8.06 [7.52, 8.59] | 8.07<br>[7.52, 8.56] | 8.06<br>[7.50, 8.59] | 8.06 [7.50, 8.59] | 8.05<br>[7.49, 8.56] |
| Age, years | 64.00<br>[57.00, 69.00] | 64.00<br>[57.00, 69.00] | 63.00<br>[56.00, 68.00] | 64.00<br>[57.00, 69.00] | 66.00<br>[60.00, 70.00] | 64.00<br>[57.00, 69.00] | 62.00<br>[55.00, 67.00] | 62.00<br>[55.00, 68.00] | 64.00<br>[57.00, 69.00] | 65.00<br>[59.00, 70.00] |
| Male, % | 26810<br>(45.4%) | 10068<br>(51.1 %) | 10822<br>(55.0%) | 11378<br>(57.8 %) | 8445<br>(42.9 %) | 9002<br>(45.7%) | 9363<br>(47.5 %) | 10582<br>(50.6%) | 8115<br>(41.7%) | 8113<br>(43.4 %) |
| Sleep, hours, (median [IQR]) | 7.62<br>[6.89, 8.26] | 6.52<br>[5.92, 6.89] | 7.62 [7.41, 7.83] | 8.52<br>[8.26, 8.89] | 7.60<br>[6.78, 8.30] | 7.65 [6.92, 8.27] | 7.62<br>[6.94, 8.22] | 7.61<br>[6.86, 8.26] | 7.62 [6.90, 8.25] | 7.64<br>[6.90, 8.27] |
| Moderate to vigorous physical activity, (median [IQR]) | 31.26<br>[18.52, 49.09] | 30.17<br>[17.52, 48.19] | 33.05<br>[19.79, 51.31] | 30.60<br>[18.24, 47.79] | 14.31<br>[9.64, 18.52] | 31.26<br>[26.76, 36.26] | 58.74<br>[49.09, 74.91] | 30.57<br>[18.24, 48.31] | 31.67<br>[18.76, 49.43] | 31.60<br>[18.59, 49.64] |
| Diet quality score, (median [IQR]) | 54.29<br>[47.50, 60.00] | 53.93<br>[47.50, 60.00] | 54.01<br>[47.50, 60.00] | 54.64<br>[47.50, 60.00] | 53.93<br>[47.50, 60.00] | 54.29<br>[47.50, 60.00] | 54.29<br>[47.50, 60.00] | 45.00<br>[40.00, 47.50] | 55.00<br>[52.50, 56.79] | 62.50<br>[60.00, 67.50] |
| Light physical activity, (median [IQR]) | 102.84<br>[70.24, 158.12] | 106.72<br>[72.26, 162.46] | 105.17<br>[72.00, 162.25] | 96.81<br>[66.72, 149.53] | 83.96<br>[59.79, 118.6] | 107.43<br>[73.38, 157.72] | 130.86<br>[81.72, 195.7] | 100.81<br>[69.03, 155.55] | 103.91<br>[71.26, 158.90] | 103.98<br>[70.53, 159.82] |
| Discretionary screen time, hours, (median [IQR]) | 3.50<br>[2.50, 5.00] | 4.00<br>[2.50, 5.00] | 3.50 [2.50, 5.00] | 3.50<br>[2.50, 5.00] | 4.00<br>[3.00, 5.00] | 3.50 [2.50, 5.00] | 3.00<br>[2.00, 4.50] | 4.00<br>[2.50, 5.00] | 3.50 [2.50, 5.00] | 3.50<br>[2.50, 5.00] |
| Smoking history, % | — | — | — | — | — | — | — | — | — | — |
| Never | 34024<br>(57.6%) | 10910<br>(55.4 %) | 11508<br>(58.4%) | 11606<br>(58.9 %) | 10781<br>(54.7 %) | 11462<br>(58.2%) | 11781<br>(59.8 %) | 11928<br>(57.0%) | 11385<br>(58.5%) | 10711<br>(57.3 %) |
| Former | 21361<br>(36.2%) | 7320<br>(37.2 %) | 7026<br>(35.7%) | 7015<br>(35.6 %) | 7390<br>(37.5 %) | 7076<br>(35.9%) | 6895<br>(35.0 %) | 7236<br>(34.6%) | 7039<br>(36.2%) | 7086<br>(37.9 %) |
| Current | 3693<br>(6.3%) | 1463<br>(7.4%) | 1158<br>(5.9%) | 1072<br>(5.4%) | 1522<br>(7.7%) | 1154<br>(5.9%) | 1017<br>(5.2%) | 1754<br>(8.4%) | 1040<br>(5.3%) | 899<br>(4.8%) |
| Alcohol consumption, units (median [IQR]) | 9.75<br>[1.70, 19.50] | 9.52<br>[0.97, 19.50] | 9.75 [2.44, 19.50] | 9.75<br>[2.27, 19.57] | 8.77<br>[0.62, 18.84] | 9.75 [2.27, 19.50] | 10.71<br>[3.20, 20.52] | 9.75<br>[1.22, 20.13] | 9.75 [2.44, 19.50] | 9.75<br>[1.94, 19.50] |
| Townsend Deprivation Index, (median [IQR]) | -2.49 [-3.84, -0.26] | -2.33<br>[-3.76, 0.13] | -2.50 [-3.83, -0.34] | -2.64<br>[-3.91, -0.59] | -2.41<br>[-3.78, -0.12] | -2.54 [-3.88, -0.34] | -2.52<br>[-3.85, -0.31] | -2.44 [-3.81, -0.09] | -2.52 [-3.87, -0.40] | -2.52<br>[-3.84, -0.33] |
| Education, % | — | — | — | — | — | — | — | — | — | — |
| College/University | 7707<br>(13.0%) | 2615<br>(13.3 %) | 2533<br>(12.9%) | 2559<br>(13.0 %) | 2523<br>(12.8 %) | 2605<br>(13.2%) | 2579<br>(13.1 %) | 2740<br>(13.1%) | 2636<br>(13.5%) | 2331<br>(12.5 %) |
| A/AS | 25619<br>(43.4%) | 8642<br>(43.9 %) | 8882<br>(45.1%) | 8095<br>(41.1 %) | 8095<br>(41.1 %) | 8725<br>(44.3%) | 8799<br>(44.7 %) | 8574<br>(41.0%) | 8780<br>(45.1%) | 8265<br>(44.2 %) |
| O levels | 3262<br>(5.5%) | 1090<br>(5.5%) | 1065<br>(5.4%) | 1107<br>(5.6%) | 1145<br>(5.8%) | 1097<br>(5.6%) | 1020<br>(5.2%) | 1264<br>(6.0%) | 989<br>(5.1%) | 1009<br>(5.4%) |
| **CSE** | 12028<br>(20.4%) | 3897<br>(19.8<br>%) | 3923<br>(19.9%) | 4208<br>(21.4<br>%) | 4104<br>(20.8<br>%) | 3982<br>(20.2%) | 3942<br>(20.0<br>%) | 4422<br>(21.1%) | 3896<br>(20.0%) | 3710<br>(19.8<br>%) |
| **NVQ/HND/HNC** | 2216<br>(3.8%) | 716<br>(3.6%) | 714<br>(3.6%) | 786<br>(4.0%) | 642<br>(3.3%) | 685<br>(3.5%) | 889<br>(4.5%) | 943<br>(4.5%) | 669<br>(3.4%) | 604<br>(3.2%) |
| **Other** | 8246<br>(14.0%) | 2733<br>(13.9<br>%) | 2575<br>(13.1%) | 2938<br>(14.9<br>%) | 3184<br>(16.2<br>%) | 2598<br>(13.2%) | 2464<br>(12.5<br>%) | 2975<br>(14.2%) | 2494<br>(12.8%) | 2777<br>(14.9<br>%) |
| **Parental history of CVD, %** | 33120<br>(56.1%) | 11074<br>(56.2<br>%) | 10976<br>(55.7%) | 11070<br>(56.2<br>%) | 11510<br>(58.4<br>%) | 11004<br>(55.9%) | 10606<br>(53.9<br>%) | 11144<br>(53.3%) | 11050<br>(56.8%) | 10926<br>(58.4<br>%) |
| **Parental history of cancer, %** | 18549<br>(31.4%) | 6177<br>(31.4<br>%) | 6150<br>(31.2%) | 6222<br>(31.6<br>%) | 6162<br>(31.3<br>%) | 6252<br>(31.7%) | 6135<br>(31.2<br>%) | 6509<br>(31.1%) | 6167<br>(31.7%) | 5873<br>(31.4<br>%) |
| **Previous CVD, %** | 5678<br>(9.6%) | 2132<br>(10.8<br>%) | 1754<br>(8.9%) | 1792<br>(9.1%) | 2668<br>(13.5<br>%) | 1732<br>(8.8%) | 1278<br>(6.5%) | 1971<br>(9.4%) | 1796<br>(9.2%) | 1911<br>(10.2<br>%) |
| **Previous Cancer, %** | 5145<br>(8.7%) | 1676<br>(8.5%) | 1672<br>(8.5%) | 1797<br>(9.1%) | 2036<br>(10.3<br>%) | 1715<br>(8.7%) | 1394<br>(7.1%) | 1709<br>(8.2%) | 1737<br>(8.9%) | 1699<br>(9.1%) |
| **Ethnicity, %** | — | — | — | — | — | — | — | — | — | — |
| **Other** | 3565<br>(6.0%) | 1552<br>(7.9%) | 1115<br>(5.7%) | 898<br>(4.6%) | 1160<br>(5.9%) | 1162<br>(5.9%) | 1243<br>(6.3%) | 1257<br>(6.0%) | 1186<br>(6.1%) | 1122<br>(6.0%) |
| **White** | 55513<br>(94.0%) | 18141<br>(92.1<br>%) | 18577<br>(94.3%) | 18795<br>(95.4<br>%) | 18533<br>(94.1<br>%) | 18530<br>(94.1%) | 18450<br>(93.7<br>%) | 19661<br>(94.0%) | 18278<br>(93.9%) | 17574<br>(94.0<br>%) |
| **Medication Use, %** | — | — | — | — | — | — | — | — | — | — |
| **Cholesterol** | 9130<br>(15.5%) | 3426<br>(17.4<br>%) | 2756<br>(14.0%) | 2948<br>(15.0<br>%) | 4200<br>(21.3<br>%) | 2870<br>(14.6%) | 2060<br>(10.5<br>%) | 2835<br>(13.6%) | 2855<br>(14.7%) | 3440<br>(18.4<br>%) |
| **Blood pressure** | 5384<br>(9.1%) | 1997<br>(10.1<br>%) | 1680<br>(8.5%) | 1707<br>(8.7%) | 2287<br>(11.6<br>%) | 1723<br>(8.7%) | 1374<br>(7.0%) | 1782<br>(8.5%) | 1749<br>(9.0%) | 1853<br>(9.9%) |
| **Insulin** | 72<br>(0.1%) | 21<br>(0.1%) | 29 (0.1%) | 22<br>(0.1%) | 22<br>(0.1%) | 27 (0.1%) | 23<br>(0.1%) | 23<br>(0.1%) | 18 (0.1%) | 31<br>(0.2%) |
| **Frailty Index >3, %** | 196<br>(0.3%) | 95<br>(0.5%) | 51 (0.3%) | 50<br>(0.3%) | 142<br>(0.8%) | 24 (0.1%) | 30<br>(0.2%) | 91<br>(0.5%) | 53 (0.3%) | 52<br>(0.3%) |
| **Self-rated health, %** | — | — | — | — | — | — | — | — | — | — |
| **Excellent** | 12827<br>(21.8%) | 3930<br>(20.0<br>%) | 4534<br>(23.1%) | 4363<br>(22.2<br>%) | 3148<br>(16.0<br>%) | 4382<br>(22.3%) | 5297<br>(26.9<br>%) | 4138<br>(19.8%) | 4322<br>(22.2%) | 4367<br>(23.4<br>%) |
| **Good** | 35514<br>(60.2%) | 11533<br>(58.7<br>%) | 11970<br>(60.9%) | 12011<br>(61.1<br>%) | 11583<br>(58.9<br>%) | 12099<br>(61.5%) | 11832<br>(60.2<br>%) | 12421<br>(59.5%) | 11897<br>(61.2%) | 11196<br>(60.0<br>%) |
| **Fair** | 9175<br>(15.6%) | 3563<br>(18.1<br>%) | 2774<br>(14.1%) | 2838<br>(14.4<br>%) | 4083<br>(20.8<br>%) | 2803<br>(14.3%) | 2289<br>(11.6<br>%) | 3654<br>(17.5%) | 2814<br>(14.5%) | 2707<br>(14.5<br>%) |
| **Poor** | 1458<br>(2.5%) | 634<br>(3.2%) | 378<br>(1.9%) | 446<br>(2.3%) | 844<br>(4.3%) | 374<br>(1.9%) | 240<br>(1.2%) | 660<br>(3.2%) | 394<br>(2.0%) | 404<br>(2.2%) |
| **Body mass index, kg/m<sup>2</sup>** | 26.10<br>[23.60,<br>29.00] | 26.70<br>[24.10,<br>29.90] | 25.80<br>[23.50,<br>28.70] | 25.70<br>[23.30,<br>28.50] | 27.20<br>[24.50,<br>30.50] | 26.00<br>[23.60,<br>28.80] | 25.20<br>[23.00,<br>27.70] | 26.40<br>[23.90,<br>29.40] | 26.00<br>[23.60,<br>28.90] | 25.80<br>[23.40,<br>28.70] |

Values represent mean (SD) unless stated otherwise stated

### Dose-response of individual SPAN behaviours with all-cause mortality

The dose-response relationship for sleep, MVPA, and DQS individually is presented in **Supplemental Figure 2**. MVPA showed an L-shaped relationship with all-cause mortality risk. When exploring physical activity in isolation compared to the 5^th^ percentile, an additional 2 min/day MVPA was associated with approximately 10% reduction in risk (HR: 0.90; 95% CI: 0.90, 0.92). Sleep revealed a J-shaped relationship where the beneficial associations of additional sleep plateaued at 7.3 hours/day. An additional 24 min/day of sleep was associated with approximately 10% risk reduction (HR: 0.91; 95% CI: 0.88, 0.93). DQS revealed a subtle and not statistically significant dose-response relationship with mortality.

### Combined SPAN associations and all-cause mortality risk

The combined association of sleep, physical activity, and nutrition with risk of all-cause mortality on the hazard ratio scale is shown in **Figure 1**. MVPA contributed most to the gradient in all-cause mortality risk followed by sleep duration and DQS. Compared to the lowest tertile for all three exposures, low MVPA combined with moderate sleep and high DQS corresponded to a 20% lower risk for all-cause mortality (HR: 0.80; 95%CI: 0.70, 0.92). Comparatively, a combination of moderate MVPA, moderate sleep, and moderate DQS was associated with a 47% reduction in risk (HR: 0.53; 95%CI: 0.46, 0.64). The highest risk reduction was associated with high MVPA, moderate sleep, and high DQS which corresponded to a 55% lower risk for all-cause mortality (HR: 0.45; 95%CI: 0.37, 0.53).

The results for combined association of sleep, physical activity, and nutrition for absolute risk of mortality are shown in **Supplemental Figure 3**. The absolute risk as incidence per 100,000 person years (PY) was highest in the low sleep, low MVPA, and low DQS tertiles (Absolute Risk: 45.61 per 100,00 PY; 95%CI: 40.10, 51.88). The lowest absolute risk was associated with a combination of high MVPA, moderate sleep duration, and high DQS (Absolute Risk: 20.18 per 100,00 PY; 95%CI: 17.41, 23.39).

### Minimal variations across the three SPAN behaviours and all-cause mortality risk

**Table 2** presents the concurrent improvements in sleep, physical activity, and nutrition associated with different levels of all-cause mortality risk reductions (up to 70%). The first rows indicate the minimal differences in the combined SPAN behaviours associated with clinically relevant risk reductions. For example, compared to the combined SPAN reference point (5^th^ percentile of all SPAN behaviours, i.e., the theoretically least healthy combination), an additional 15 mins/day of sleep, 1.6 min/day MVPA, and increasing DQS by 5 points was associated with a 10% lower risk of all-cause mortality (HR: 0.90; 95% CI: 0.88, 0.93). An increase of 75 mins/day of sleep, 12.5 mins/day MVPA, and 25 DQS points corresponds to a 50% lower risk of all-cause mortality (HR: 0.50; 95% CI: 0.44, 0.58). For context, a 5-point increase in DQS corresponds to reducing processed meat consumption by half a serving weekly, or cutting back on refined grains such as bread, biscuits, and cereal by one serving per week. An increase of 25 DQS points corresponds to multiple dietary changes such as consuming a 1/3 cup per day of cooked vegetables, reducing refined grains intake by two servings per week, and eliminating consumption of sugar-sweetened beverages. The concurrent combinations of SPAN exposures and corresponding all-cause mortality risk is shown as a heat map correlogram in **Figure 2**. The absolute risk plot (**Supplemental Figure 4)** showed that risk reduction is more evenly distributed across differences in MVPA, sleep, and DQS.

**Figure 2:**
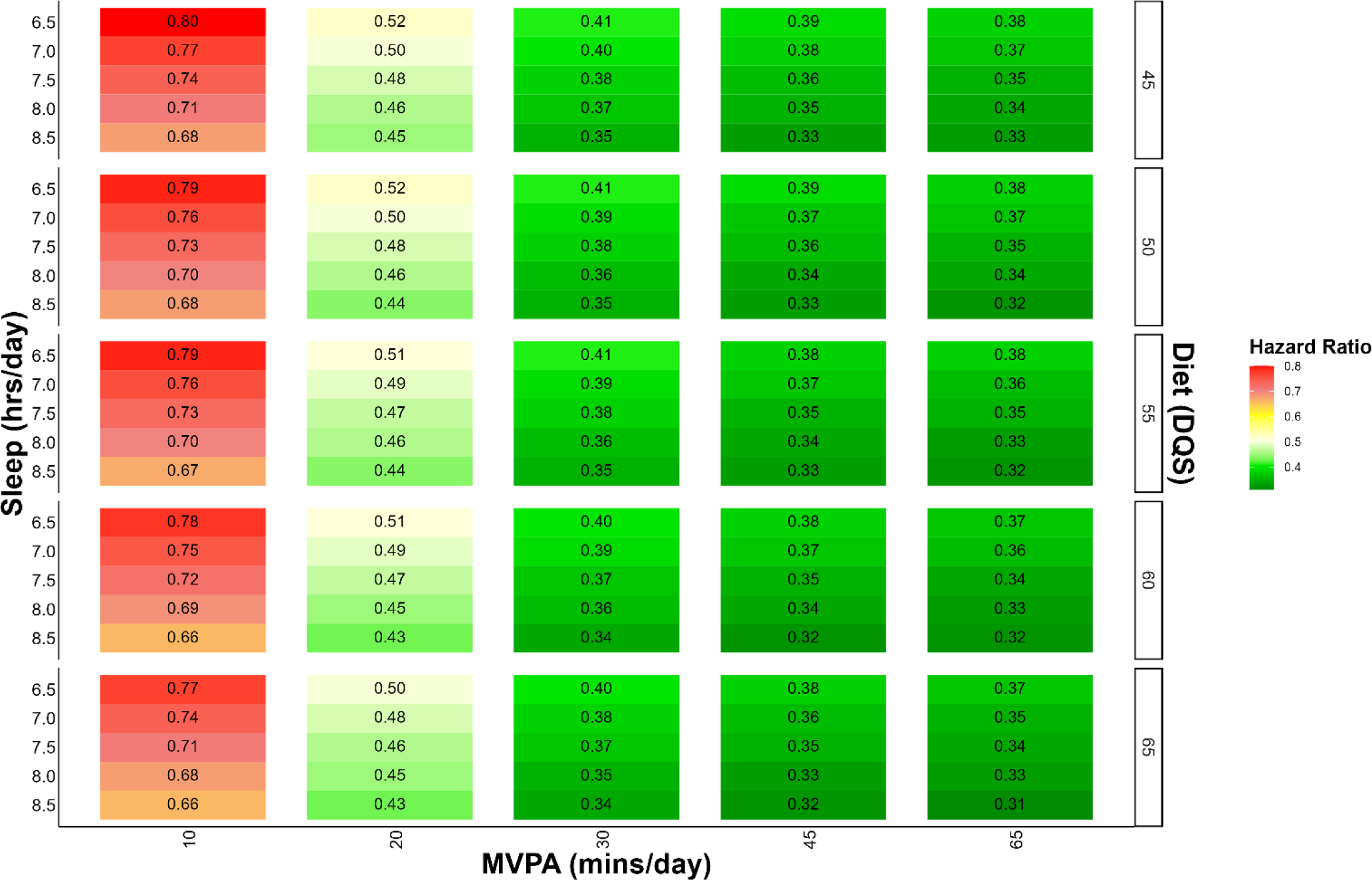
All-cause mortality risk associated with concurrent variations in sleep, MVPA, and dietary quality score The correlogram displays changes in SPAN and corresponding mortality risk with the reference being the 5^th^ percentile of sleep (5.5 hours/day), physical activity (7.3 mins/day), and nutrition (36.9 DQS). Moderate-Vigorous Physical Activity (MVPA); Diet Quality Score (DQS). Model is adjusted for age, sex, ethnicity, smoking, education, Townsend deprivation index, alcohol, discretionary screen time (time spent watching TV or using the computer outside of work), light intensity physical activity, medication (blood pressure, insulin, and cholesterol), previous diagnosis of major CVD (defined as disease of the circulatory system, arteries, and lymph, excluding hypertension), previous diagnosis of cancer, and familial history of CVD and cancer.

**Table 2:**
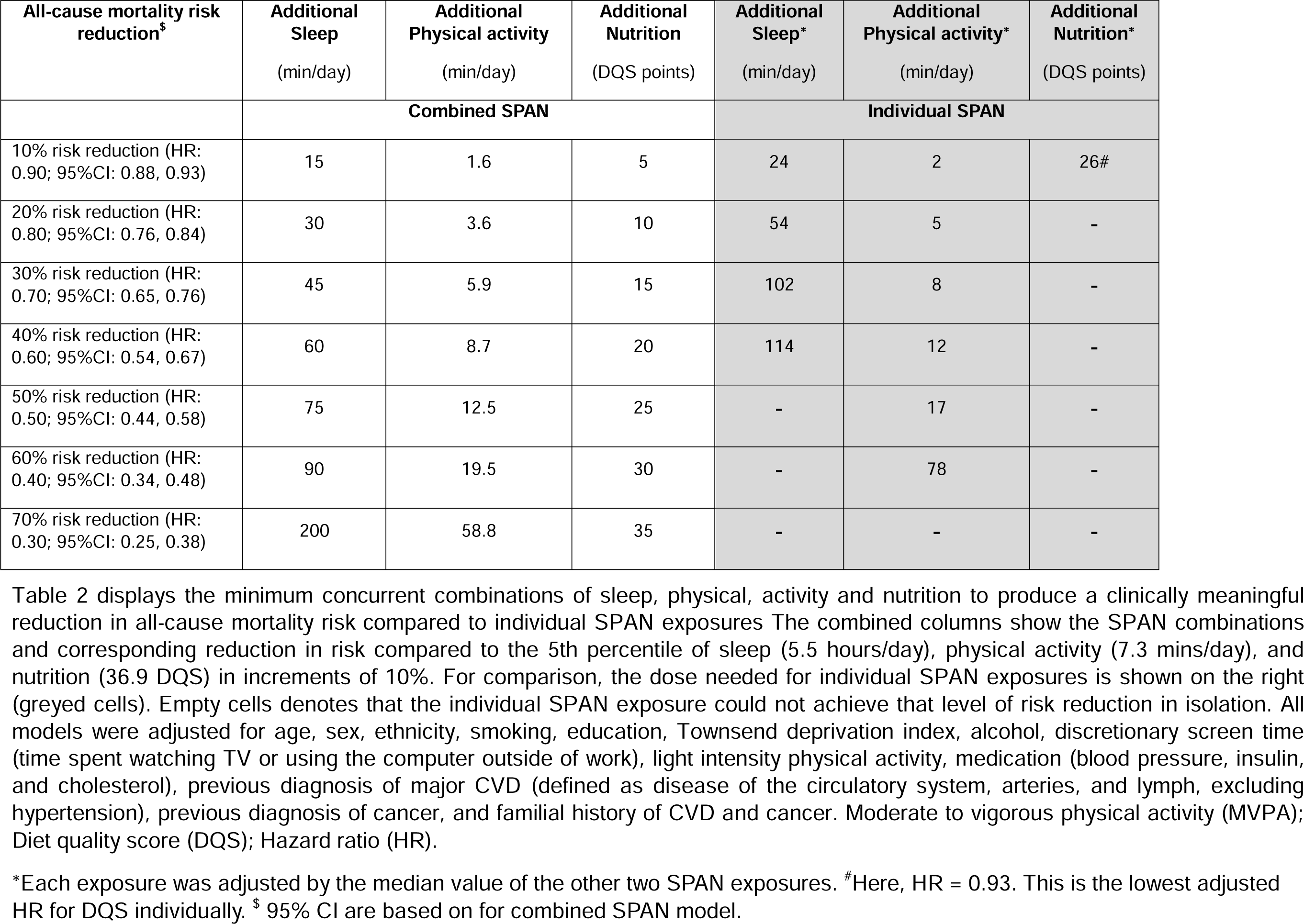
Concurrent variations in sleep, physical, and nutrition associated with increments of all-cause mortality risk reduction compared to 5^th^ percentile of SPAN exposures.

### Sensitivity Analyses

Excluding participants with poor health and adjusting for BMI did not materially influence the results **(Supplemental Figure 5-7)**.

## DISCUSSION

### Main Findings

This is the first combined sleep, physical activity, and nutrition study to estimate minimal concurrent incremental variations across the three behaviours that are associated with meaningful reductions in all-cause mortality risk. We utilised a novel multi-exposure approach and data from the wearables study of the UK Biobank for sleep and physical activity measurements [25–27, 32]. Our findings reveal that the combination of sleep, physical activity, and nutrition yield synergistic effects where the combination of high MVPA, moderate sleep, and high DQS corresponded to a 55% lower risk for all-cause mortality compared to the reference group. Notably, even modest changes (an additional 15 mins/day of sleep, 1.6 min/day MVPA, and 5 DQS points) were associated with the minimum clinically meaningful difference in all-cause mortality risk of 10%.

While a substantial body of research highlights the pairwise effects of physical activity, nutrition, and sleep [11, 12, 20, 36–38], limited research exists that explores all three factors in combination with health outcomes, particularly the combined incremental benefits on mortality. Previous joint analyses of self-reported physical activity and sleep with all-cause mortality have also shown that poor sleep can amplify the effect of physical inactivity [20]. Notably, the risk reduction for physical activity was significantly more pronounced in the present study compared to the aforementioned joint-analyses in the UK Biobank. This may have resulted from the use of objectively measured physical activity in the present study, which compared to self-report methods, has shown a threefold magnitude of benefit for all-cause mortality risk reduction [39].

Together, these results strengthen the importance of exploring the effects of lifestyle behaviours using objectively measured data.

Ding et al. previously examined the relationship between lifestyle risk behaviours and mortality risk in a group of 231,048 participants from the Australian 45 and Up Study [40]. This study revealed that individuals with short sleep duration, physical inactivity, and poor diet had a 49% increased risk of all-cause mortality compared to the referent group. However, this study relied on dichotomized self-reported data, which is subject to under and over-reporting while also lacking more detailed insights required for public health recommendations. Other work from the HUNT Study in Norway (n = 36,911), demonstrated that short sleep duration, physical inactivity and poor diet, combined with lack of social participation, led to an over two-fold increase in risk of all-cause mortality (HR: 2.26; 95% CI: 1.91-2.69) [41]. Importantly, both studies recognised that, of the three SPAN behaviours, physical inactivity was both among the most prevalent and influential risk factors.

In the present study, physical activity displayed the greatest overall effect on the risk of all-cause mortality compared to the other SPAN behaviours. This was followed by sleep which displayed a general U-shape risk curve, and the lowest overall effect was observed with diet quality. The individual associations of sleep, MVPA, and DQS, required a substantially higher dose to elicit the same risk reduction compared to combined SPAN changes. For a 10% reduction in risk, this corresponded to 60% more sleep (24 min/day), 25% more MVPA (2 min/day) and diet alone, as measured using the chosen DQS, was unable to reach this level of risk reduction in isolation. In comparison, when considering all SPAN behaviours in combination, a minimum increase of only 15 min/day for sleep, 1.6 min/day MVPA, and 5 DQS was sufficient to achieve this goal. For diet quality, this translates into a relatively modest difference such as consuming an additional 1.5 pieces of fresh fruit per day, increasing vegetable intake by 1/3 cup cooked vegetables per day, or adding one serving of fatty fish per week. This study emphasizes that combined doses of SPAN behaviours act synergistically where collective effects were associated with a significantly greater impact on all-cause mortality risk reduction than its individual components. Importantly, these results highlight that interventions addressing individual SPAN behaviours may be missing a powerful and more sustainable intervention option for reducing mortality risk.

Lifestyle risk factors often co-occur and ultimately form larger behavioral patterns that shape social groups and communities [42]. From a clinical standpoint, our results offer tangible guidance to assist individuals and practitioners in identifying the most feasible behavioural lifestyle changes. This is a crucial point for reducing the risk of noncommunicable disease and mortality considering the multiple barriers to major SPAN improvements such as inability to find time or motivation for leisure time physical exercise, limited access to healthy foods and cooking skills, or unavoidable disruptions to sleep patterns. From a public health standpoint, this evidence offers the granularity needed for providing feasible lifestyle recommendations that can significantly reduce all-cause mortality risk.

### Strengths and Limitations

A key strength of this study is the novel analytic approach estimating changes across multiple influential lifestyle behaviours corresponding to different degrees of risk reduction. This has the advantage of allowing the exploration of both combined effects and the relative influence of specific lifestyle modification components. This is of particular importance considering the well-established co-dependencies between each of the SPAN exposures [11, 13, 16, 20, 21, 40, 43]. The study is further strengthened due to the device-based measurement of both sleep and physical activity. By utilizing wearable devices for data collection, the study effectively mitigates the common pitfalls associated with self-reported data, such as under-reporting or over-reporting.

A notable limitation of the study is the reliance on subjective self-reported data for dietary intake given the absence of objectively measured dietary data in the UK Biobank. This introduces a potential misalignment in the data quality for the SPAN exposures. Such discrepancies can lead to disproportionate measurement error, which might have influenced the estimated effect size for diet quality. Additionally, dietary data was collected at baseline (2006-2009) while device measured sleep and physical activity was collected several years later (2013-2015). This temporal difference in data collection introduces the possibility that changes in lifestyle factors over time may have influenced the observed associations. The DQS may also not capture some aspects of diet that impact all-cause mortality such as total calorie and specific amino acid intake that have been shown to deeply influence average and maximal lifespan [7]. From a sleep perspective, this study explored sleep duration to provide a more interpretable outcome for behavioural change. However, other aspects of sleep such as regularity, timing, and quality of sleep play are important for health outcomes and should be considered in future research. Despite the wide breadth of confounders used, it cannot be ruled out that some degree of unmeasured confounding may be present in this analysis. Finally, this study did not examine the required duration of sustained behavioral changes for achieving clinically significant risk reduction. Further research is essential to understand the sustainability of SPAN behaviour modifications and the potential influence duration may have on these associations.

## CONCLUSIONS

In conclusion, our study underscores the significance of incremental combined positive lifestyle changes. Even modest collective improvements, such as a 15 mins/day increase in sleep, 1.6 min/day higher MVPA, and higher 5 DQS points, were associated with a clinically meaningful 10% reduction in the risk of all-cause mortality. Our findings support the implementation of clinical and educational strategies, leveraging advanced wearables and metabolic sensors technology, aimed at encouraging small incremental enhancements in sleep, physical activity, and nutrition across broad segments of the population. Further research is needed to substantiate the potential advantage of this novel approach, which is currently theoretical in nature.

## Acknowledgements

This research has been conducted using the UK Biobank resource under application number 25813. The authors would like to thank all the participants and professionals contributing to the UK Biobank. All information and materials in the manuscript are original and have not been submitted for publication elsewhere.

## Data availability Statement

The data that support the findings of this study are available from the UK Biobank, but restrictions apply to the availability of these data, which were used under license for the current study, and so are not publicly available. Data are however available from the authors upon reasonable request and with the permission of the UK Biobank.

