## Supplemental Material for "Minimum changes in sleep, physical activity, and nutrition associated with clinically important reductions in all-cause mortality risk: a prospective cohort study"

**Affiliations:**

**Word Count**: Main text: 3,418; Abstract: 348; Tables/Figures: 4; Supplemental Material: 9 items.

**Keywords**: Sleep, nutrition, physical activity, mortality, cohort studies

#### ABSTRACT

**Background:** Sleep, physical activity, and nutrition (SPAN) are crucial modifiable factors for health, yet most research has examined them independently rather than exploring their combined and incremental impact on disease risk and mortality.

**Funding:** This study is funded by an Australian National Health and Medical Research Council (NHMRC) Investigator Grant (APP1194510). The funder had no specific role in any of the following study aspects: the design and conduct of the study; collection, management, analysis, and interpretation of the data; preparation, review, or approval of the manuscript; and decision to submit the manuscript for publication. L.F. is supported by grants from the Australian NHMRC Investigator Grant (APP1177797) and Australian Youth and Health Foundation. D.D. is supported by the Australian Research Council DECRA program (DE230101174). B.D.P.C. is supported by the Government of Andalusia (Spain), Research Talent Recruitment Program (EMERGIA 2020/00158).

**Acknowledgements:** This research has been conducted using the UK Biobank resource under application number 25813. The authors would like to thank all the participants and professionals contributing to the UK Biobank. All information and materials in the manuscript are original and have not been submitted for publication elsewhere.

**Data availability Statement**: The data that support the findings of this study are available from the UK Biobank, but restrictions apply to the availability of these data, which were used under license for the current study, and so are not publicly available. Data are however available from the authors upon reasonable request and with the permission of the UK Biobank.

**Conflicts of Interest:** None.

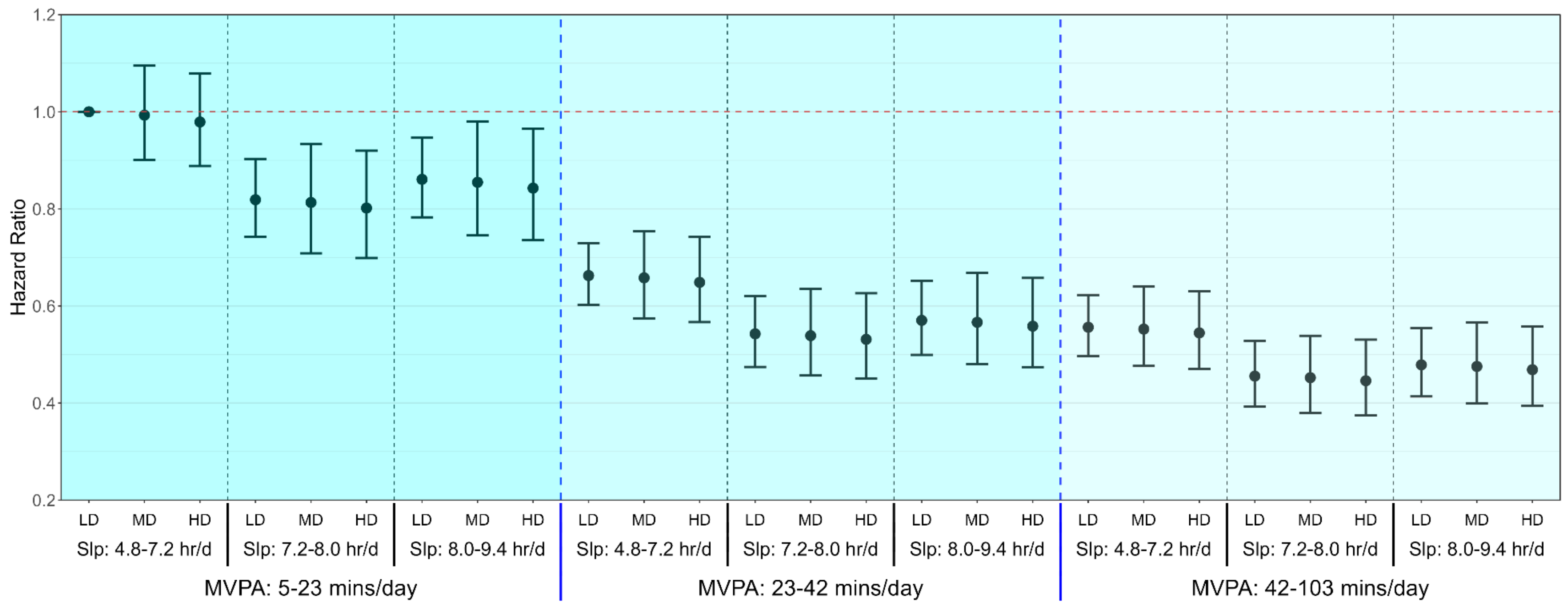


**Figure 1:** Association of sleep, MVPA, and diet quality with all-cause mortality

**Legend:** Model is adjusted for age, sex, ethnicity, smoking, education, Townsend deprivation index, alcohol, discretionary screen time (time spent watching TV or using the computer outside of work), light intensity physical activity, medication (blood pressure, insulin, and cholesterol), previous diagnosis of major CVD (defined as disease of the circulatory system, arteries, and lymph, excluding hypertension), previous diagnosis of cancer, and familial history of CVD and cancer. Dashed blue lines separate tertiles MVPA and dashed black lines separate tertiles of sleep. Moderate-Vigorous Physical Activity (MVPA); Low Diet Quality (LD); Medium Diet Quality (MD); High Diet Quality (HD).

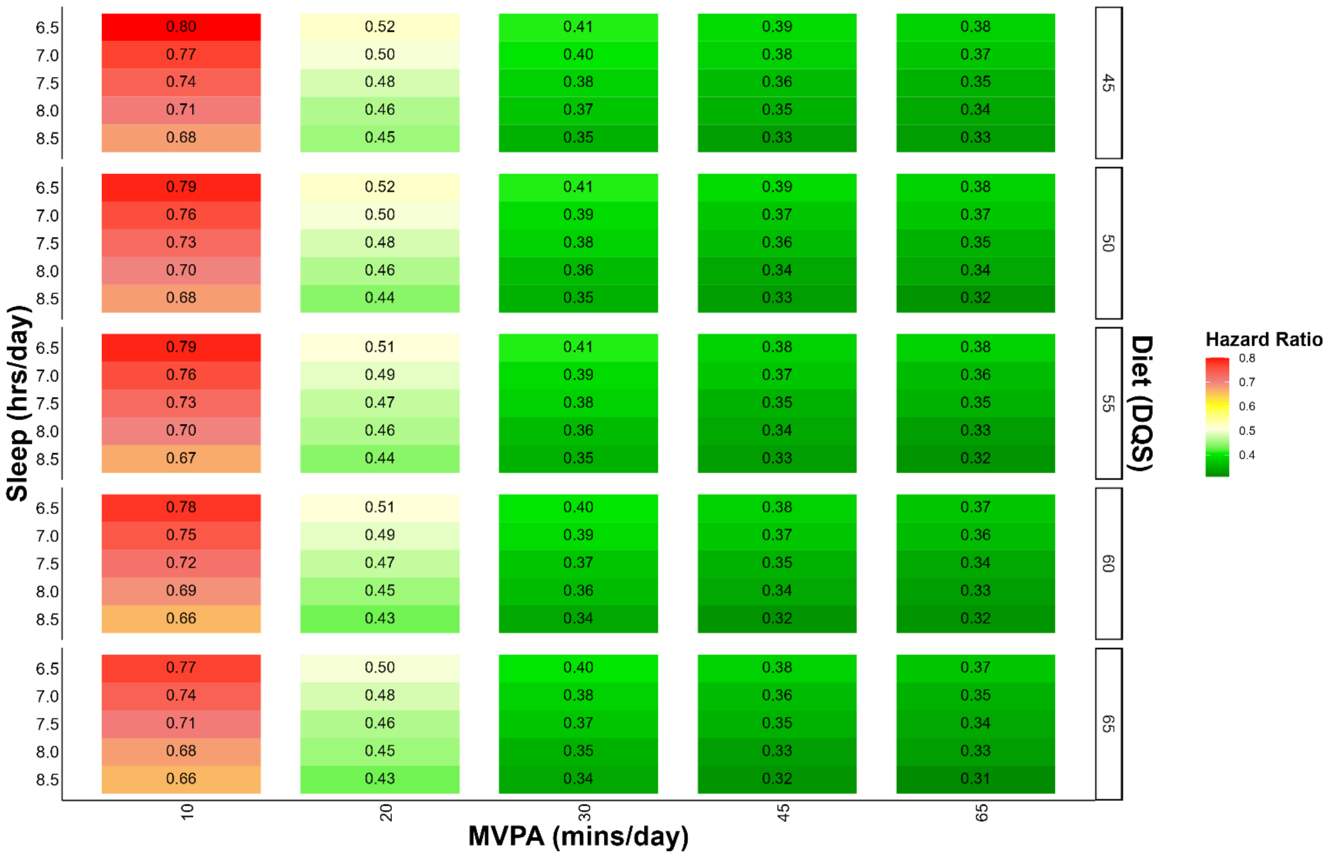


**Figure 2:** All-cause mortality risk associated with concurrent variations in sleep, MVPA, and dietary quality score

**Legend:** The correlogram displays changes in SPAN and corresponding mortality risk with the reference being the 5$^{th}$ percentile of sleep (5.5 hours/day), physical activity (7.3 mins/day), and nutrition (36.9 DQS). Moderate-Vigorous Physical Activity (MVPA); Diet Quality Score (DQS). Model is adjusted for age, sex, ethnicity, smoking, education, Townsend deprivation index, alcohol, discretionary screen time (time spent watching TV or using the computer outside of work), light intensity physical activity, medication (blood pressure, insulin, and cholesterol), previous diagnosis of major CVD (defined as disease of the circulatory system, arteries, and lymph, excluding hypertension), previous diagnosis of cancer, and familial history of CVD and cancer.

**Table 1:** Participant characteristics

| | Overall | Sleep | | | Physical activity | | | Nutrition | | |
|---|---|---|---|---|---|---|---|---|---|---|
| | | Low | Moderate | High | Low | Moderate | High | Low | Moderate | High |
| **Sample** | 59078 | 19693 | 19692 | 19693 | 19693 | 19692 | 19693 | 20918 | 19464 | 18696 |
| **Events, n** | 2458 | 980 | 698 | 780 | 1276 | 693 | 489 | 856 | 783 | 819 |
| **Follow up, years** | 8.06 [7.49, 8.59] | 8.05 [7.48, 8.56] | 8.07 [7.51, 8.59] | 8.05 [7.51, 8.59] | 8.05 [7.46, 8.59] | 8.06 [7.52, 8.59] | 8.07 [7.52, 8.56] | 8.06 [7.50, 8.59] | 8.06 [7.50, 8.59] | 8.05 [7.49, 8.56] |
| **Age, years** | 64.00 [57.00, 69.00] | 64.00 [57.00, 69.00] | 63.00 [56.00, 68.00] | 64.00 [57.00, 69.00] | 66.00 [60.00, 70.00] | 64.00 [57.00, 69.00] | 62.00 [55.00, 67.00] | 62.00 [55.00, 68.00] | 64.00 [57.00, 69.00] | 65.00 [59.00, 70.00] |
| **Male, %** | 26810 (45.4%) | 10068 (51.1%) | 10822 (55.0%) | 11378 (57.8%) | 8445 (42.9%) | 9002 (45.7%) | 9363 (47.5%) | 10582 (50.6%) | 8115 (41.7%) | 8113 (43.4%) |
| **Sleep, hours, (median [IQR])** | 7.62 [6.89, 8.26] | 6.52 [5.92, 6.89] | 7.62 [7.41, 7.83] | 8.52 [8.26, 8.89] | 7.60 [6.78, 8.30] | 7.65 [6.92, 8.27] | 7.62 [6.94, 8.22] | 7.61 [6.86, 8.26] | 7.62 [6.90, 8.25] | 7.64 [6.90, 8.27] |
| **Moderate to vigorous physical activity, (median [IQR])** | 31.26 [18.52, 49.09] | 30.17 [17.52, 48.19] | 33.05 [19.79, 51.31] | 30.60 [18.24, 47.79] | 14.31 [9.64, 18.52] | 31.26 [26.76, 36.26] | 58.74 [49.09, 74.91] | 30.57 [18.24, 48.31] | 31.67 [18.76, 49.43] | 31.60 [18.59, 49.64] |
| **Diet quality score, (median [IQR])** | 54.29 [47.50, 60.00] | 53.93 [47.50, 60.00] | 54.01 [47.50, 60.00] | 54.64 [47.50, 60.00] | 53.93 [47.50, 60.00] | 54.29 [47.50, 60.00] | 54.29 [47.50, 60.00] | 45.00 [40.00, 47.50] | 55.00 [52.50, 56.79] | 62.50 [60.00, 67.50] |
| **Light physical activity, (median [IQR])** | 102.84 [70.24, 158.12] | 106.72 [72.26, 162.46] | 105.17 [72.00, 162.25] | 96.81 [66.72, 149.53] | 83.96 [59.79, 118.6] | 107.43 [73.38, 157.72] | 130.86 [81.72, 195.7] | 100.81 [69.03, 155.55] | 103.91 [71.26, 158.90] | 103.98 [70.53, 159.82] |
| **Discretionary screen time, hours, (median [IQR])** | 3.50 [2.50, 5.00] | 4.00 [2.50, 5.00] | 3.50 [2.50, 5.00] | 3.50 [2.50, 5.00] | 4.00 [3.00, 5.00] | 3.50 [2.50, 5.00] | 3.00 [2.00, 4.50] | 4.00 [2.50, 5.00] | 3.50 [2.50, 5.00] | 3.50 [2.50, 5.00] |
| **Smoking history, %** | — | — | — | — | — | — | — | — | — | — |
| **Never** | 34024 (57.6%) | 10910 (55.4%) | 11508 (58.4%) | 11606 (58.9%) | 10781 (54.7%) | 11462 (58.2%) | 11781 (59.8%) | 11928 (57.0%) | 11385 (58.5%) | 10711 (57.3%) |
| **Former** | 21361 (36.2%) | 7320 (37.2%) | 7026 (35.7%) | 7015 (35.6%) | 7390 (37.5%) | 7076 (35.9%) | 6895 (35.0%) | 7236 (34.6%) | 7039 (36.2%) | 7086 (37.9%) |
| **Current** | 3693 (6.3%) | 1463 (7.4%) | 1158 (5.9%) | 1072 (5.4%) | 1522 (7.7%) | 1154 (5.9%) | 1017 (5.2%) | 1754 (8.4%) | 1040 (5.3%) | 899 (4.8%) |
| **Alcohol consumption, units (median [IQR])** | 9.75 [1.70, 19.50] | 9.52 [0.97, 19.50] | 9.75 [2.44, 19.50] | 9.75 [2.27, 19.57] | 8.77 [0.62, 18.84] | 9.75 [2.27, 19.50] | 10.71 [3.20, 20.52] | 9.75 [1.22, 20.13] | 9.75 [2.44, 19.50] | 9.75 [1.94, 19.50] |
| **Townsend Deprivation Index, (median [IQR]])** | -2.49 [-3.84, -0.26] | -2.33 [-3.76, 0.13] | -2.50 [-3.83, -0.34] | -2.64 [-3.91, -0.59] | -2.41 [-3.78, -0.12] | -2.54 [-3.88, -0.34] | -2.52 [-3.85, -0.31] | -2.44 [-3.81, -0.09] | -2.52 [-3.87, -0.40] | -2.52 [-3.84, -0.33] |
| **Education, %** | — | — | — | — | — | — | — | — | — | — |
| **College/University** | 7707 (13.0%) | 2615 (13.3%) | 2533 (12.9%) | 2559 (13.0%) | 2523 (12.8%) | 2605 (13.2%) | 2579 (13.1%) | 2740 (13.1%) | 2636 (13.5%) | 2331 (12.5%) |
| **A/AS** | 25619 (43.4%) | 8642 (43.9%) | 8882 (45.1%) | 8095 (41.1%) | 8095 (41.1%) | 8725 (44.3%) | 8799 (44.7%) | 8574 (41.0%) | 8780 (45.1%) | 8265 (44.2%) |
| **O levels** | 3262 (5.5%) | 1090 (5.5%) | 1065 (5.4%) | 1107 (5.6%) | 1145 (5.8%) | 1097 (5.6%) | 1020 (5.2%) | 1264 (6.0%) | 989 (5.1%) | 1009 (5.4%) |

| | | | | | | | | | | |
|---|---|---|---|---|---|---|---|---|---|---|
| **CSE** | 12028 (20.4%) | 3897 (19.8%) | 3923 (19.9%) | 4208 (21.4%) | 4104 (20.8%) | 3982 (20.2%) | 3942 (20.0%) | 4422 (21.1%) | 3896 (20.0%) | 3710 (19.8%) |
| **NVQ/HND/HNC** | 2216 (3.8%) | 716 (3.6%) | 714 (3.6%) | 786 (4.0%) | 642 (3.3%) | 685 (3.5%) | 889 (4.5%) | 943 (4.5%) | 669 (3.4%) | 604 (3.2%) |
| **Other** | 8246 (14.0%) | 2733 (13.9%) | 2575 (13.1%) | 2938 (14.9%) | 3184 (16.2%) | 2598 (13.2%) | 2464 (12.5%) | 2975 (14.2%) | 2494 (12.8%) | 2777 (14.9%) |
| **Parental history of CVD, %** | 33120 (56.1%) | 11074 (56.2%) | 10976 (55.7%) | 11070 (56.2%) | 11510 (58.4%) | 11004 (55.9%) | 10606 (53.9%) | 11144 (53.3%) | 11050 (56.8%) | 10926 (58.4%) |
| **Parental history of cancer, %** | 18549 (31.4%) | 6177 (31.4%) | 6150 (31.2%) | 6222 (31.6%) | 6162 (31.3%) | 6252 (31.7%) | 6135 (31.2%) | 6509 (31.1%) | 6167 (31.7%) | 5873 (31.4%) |
| **Previous CVD, %** | 5678 (9.6%) | 2132 (10.8%) | 1754 (8.9%) | 1792 (9.1%) | 2668 (13.5%) | 1732 (8.8%) | 1278 (6.5%) | 1971 (9.4%) | 1796 (9.2%) | 1911 (10.2%) |
| **Previous Cancer, %** | 5145 (8.7%) | 1676 (8.5%) | 1672 (8.5%) | 1797 (9.1%) | 2036 (10.3%) | 1715 (8.7%) | 1394 (7.1%) | 1709 (8.2%) | 1737 (8.9%) | 1699 (9.1%) |
| **Ethnicity, %** | — | — | — | — | — | — | — | — | — | — |
| **Other** | 3565 (6.0%) | 1552 (7.9%) | 1115 (5.7%) | 898 (4.6%) | 1160 (5.9%) | 1162 (5.9%) | 1243 (6.3%) | 1257 (6.0%) | 1186 (6.1%) | 1122 (6.0%) |
| **White** | 55513 (94.0%) | 18141 (92.1%) | 18577 (94.3%) | 18795 (95.4%) | 18533 (94.1%) | 18530 (94.1%) | 18450 (93.7%) | 19661 (94.0%) | 18278 (93.9%) | 17574 (94.0%) |
| **Medication Use, %** | — | — | — | — | — | — | — | — | — | — |
| **Cholesterol** | 9130 (15.5%) | 3426 (17.4%) | 2756 (14.0%) | 2948 (15.0%) | 4200 (21.3%) | 2870 (14.6%) | 2060 (10.5%) | 2835 (13.6%) | 2855 (14.7%) | 3440 (18.4%) |
| **Blood pressure** | 5384 (9.1%) | 1997 (10.1%) | 1680 (8.5%) | 1707 (8.7%) | 2287 (11.6%) | 1723 (8.7%) | 1374 (7.0%) | 1782 (8.5%) | 1749 (9.0%) | 1853 (9.9%) |
| **Insulin** | 72 (0.1%) | 21 (0.1%) | 29 (0.1%) | 22 (0.1%) | 22 (0.1%) | 27 (0.1%) | 23 (0.1%) | 23 (0.1%) | 18 (0.1%) | 31 (0.2%) |
| **Frailty Index >3, %** | 196 (0.3%) | 95 (0.5%) | 51 (0.3%) | 50 (0.3%) | 142 (0.8%) | 24 (0.1%) | 30 (0.2%) | 91 (0.5%) | 53 (0.3%) | 52 (0.3%) |
| **Self-rated health, %** | — | — | — | — | — | — | — | — | — | — |
| **Excellent** | 12827 (21.8%) | 3930 (20.0%) | 4534 (23.1%) | 4363 (22.2%) | 3148 (16.0%) | 4382 (22.3%) | 5297 (26.9%) | 4138 (19.8%) | 4322 (22.2%) | 4367 (23.4%) |
| **Good** | 35514 (60.2%) | 11533 (58.7%) | 11970 (60.9%) | 12011 (61.1%) | 11583 (58.9%) | 12099 (61.5%) | 11832 (60.2%) | 12421 (59.5%) | 11897 (61.2%) | 11196 (60.0%) |
| **Fair** | 9175 (15.6%) | 3563 (18.1%) | 2774 (14.1%) | 2838 (14.4%) | 4083 (20.8%) | 2803 (14.3%) | 2289 (11.6%) | 3654 (17.5%) | 2814 (14.5%) | 2707 (14.5%) |
| **Poor** | 1458 (2.5%) | 634 (3.2%) | 378 (1.9%) | 446 (2.3%) | 844 (4.3%) | 374 (1.9%) | 240 (1.2%) | 660 (3.2%) | 394 (2.0%) | 404 (2.2%) |
| **Body mass index, kg/m$^2$** | 26.10 [23.60, 29.00] | 26.70 [24.10, 29.90] | 25.80 [23.50, 28.70] | 25.70 [23.30, 28.50] | 27.20 [24.50, 30.50] | 26.00 [23.60, 28.80] | 25.20 [23.00, 27.70] | 26.40 [23.90, 29.40] | 26.00 [23.60, 28.90] | 25.80 [23.40, 28.70] |

Values represent mean (SD) unless stated otherwise stated

**Table 2: Concurrent variations in sleep, physical, and nutrition associated with increments of all-cause mortality risk reduction compared to 5th percentile of SPAN exposures**

| All-cause mortality risk reduction$ | Additional Sleep (min/day) | Additional Physical activity (min/day) | Additional Nutrition (DQS points) | Additional Sleep* (min/day) | Additional Physical activity* (min/day) | Additional Nutrition* (DQS points) |
|---|---|---|---|---|---|---|
| | **Combined SPAN** | | | **Individual SPAN** | | |
| 10% risk reduction (HR: 0.90; 95%CI: 0.88, 0.93) | 15 | 1.6 | 5 | 24 | 2 | 26# |
| 20% risk reduction (HR: 0.80; 95%CI: 0.76, 0.84) | 30 | 3.6 | 10 | 54 | 5 | - |
| 30% risk reduction (HR: 0.70; 95%CI: 0.65, 0.76) | 45 | 5.9 | 15 | 102 | 8 | - |
| 40% risk reduction (HR: 0.60; 95%CI: 0.54, 0.67) | 60 | 8.7 | 20 | 114 | 12 | - |
| 50% risk reduction (HR: 0.50; 95%CI: 0.44, 0.58) | 75 | 12.5 | 25 | - | 17 | - |
| 60% risk reduction (HR: 0.40; 95%CI: 0.34, 0.48) | 90 | 19.5 | 30 | - | 78 | - |
| 70% risk reduction (HR: 0.30; 95%CI: 0.25, 0.38) | 200 | 58.8 | 35 | - | - | - |

Table 2 displays the minimum concurrent combinations of sleep, physical, activity and nutrition to produce a clinically meaningful reduction in all-cause mortality risk compared to individual SPAN exposures The combined columns show the SPAN combinations and corresponding reduction in risk compared to the 5th percentile of sleep (5.5 hours/day), physical activity (7.3 mins/day), and nutrition (36.9 DQS) in increments of 10%. For comparison, the dose needed for individual SPAN exposures is shown on the right (greyed cells). Empty cells denotes that the individual SPAN exposure could not achieve that level of risk reduction in isolation. All models were adjusted for age, sex, ethnicity, smoking, education, Townsend deprivation index, alcohol, discretionary screen time (time spent watching TV or using the computer outside of work), light intensity physical activity, medication (blood pressure, insulin, and cholesterol), previous diagnosis of major CVD (defined as disease of the circulatory system, arteries, and lymph, excluding hypertension), previous diagnosis of cancer, and familial history of CVD and cancer. Moderate to vigorous physical activity (MVPA); Diet quality score (DQS); Hazard ratio (HR).

*Each exposure was adjusted by the median value of the other two SPAN exposures. [#]Here, HR = 0.93. This is the lowest adjusted HR for DQS individually. [$] 95% CI are based on for combined SPAN model.

**ONLINE SUPPLEMENTARY MATERIAL**

### Minimum changes in sleep, physical activity, and nutrition associated with clinically important reductions in all-cause mortality risk: a prospective cohort study

**Table of Contents**


| Page | Item |
|---|---|
| 2 | **Supplemental Figure 1**: Flow diagram of participants in the study |
| 3 | **Supplemental Figure 2:** Dose-response association of sleep, MVPA, and diet quality score with all-cause mortality risk |
| 4 | **Supplemental Figure 3**: Association of sleep, MVPA, and diet quality with absolute all-cause mortality risk |
| 5 | **Supplemental Figure 4:** Absolute all-cause mortality risk associated with concurrent variations in sleep, MVPA, and dietary quality score |
| 6 | **Supplemental Figure 5:** Association of sleep, MVPA, and diet quality with all-cause mortality excluding individuals with poor health |
| 7 | **Supplemental Figure 6:** Association of sleep, MVPA, and diet quality with all-cause mortality with adjustment for body mass index |
| 8 | **Supplemental Table 1**: Covariate definitions |
| 9 | **Supplemental Table 2:** STROBE statement |

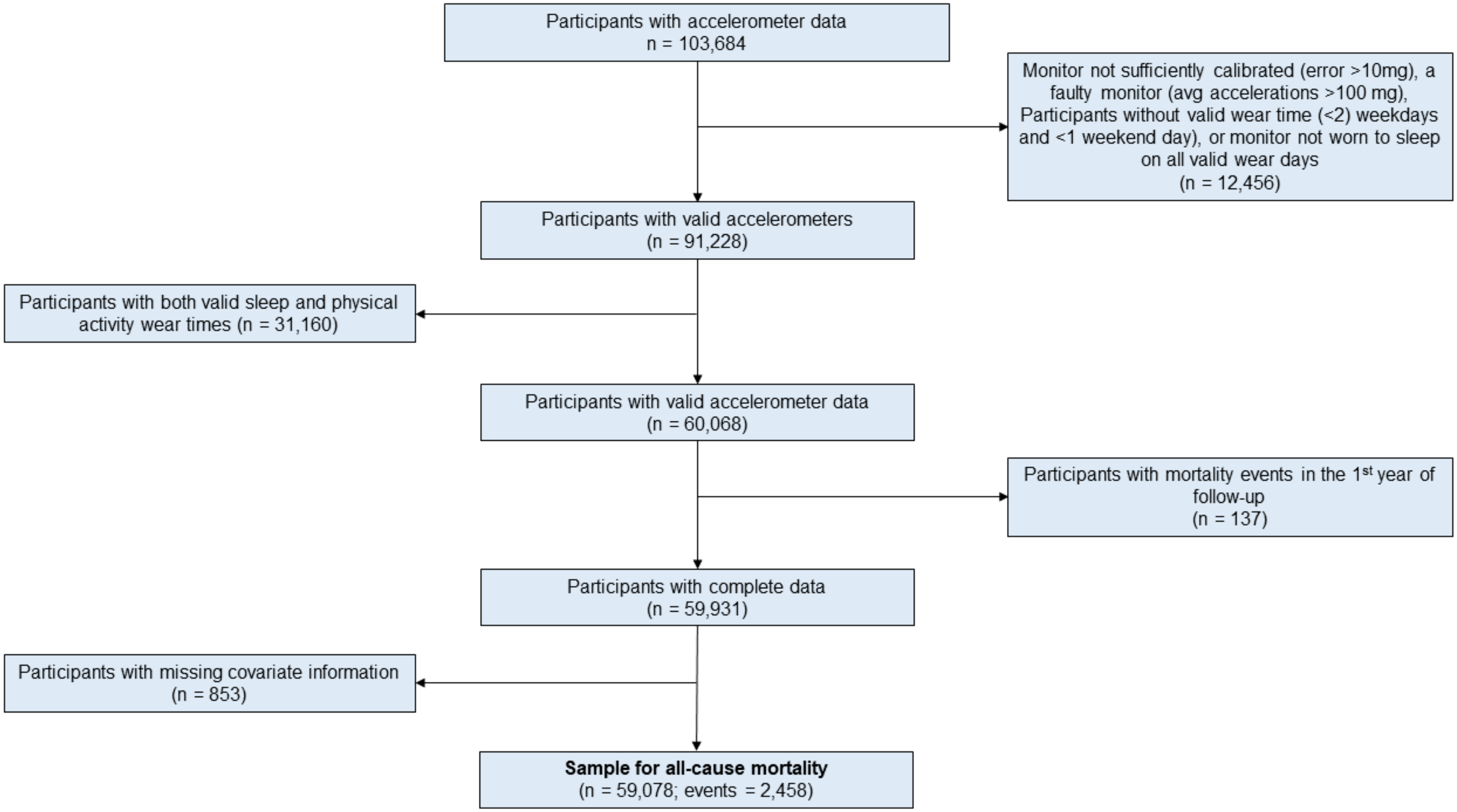


**Supplementary Figure 1.** Participant flow chart

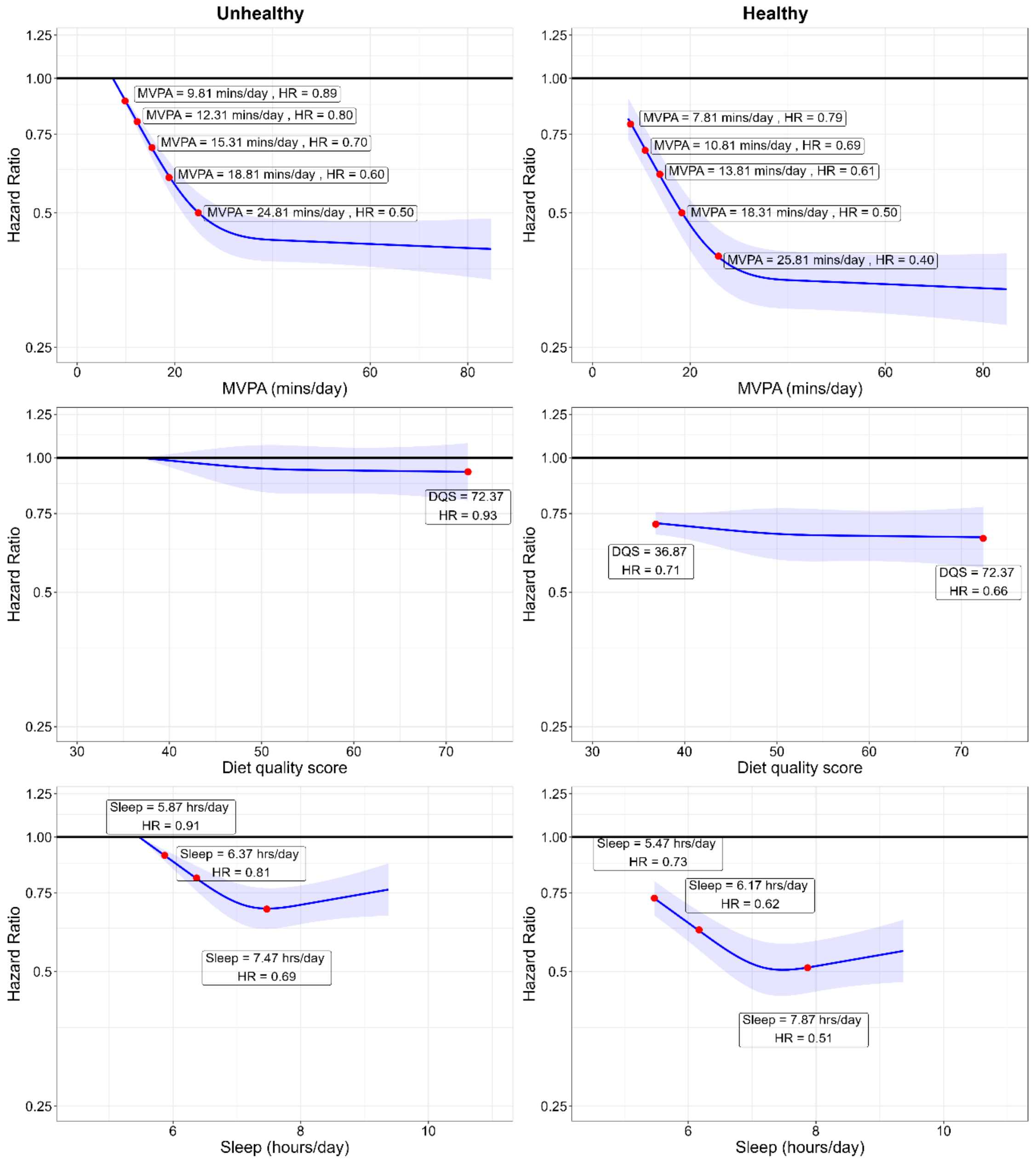


**Supplemental Figure 2:** Dose-response association of sleep, MVPA, and diet quality score with all-cause mortality risk

**Legend**: Dose-response plots for individual span exposures are shown with an 'unhealthy' reference (i.e., 5$^{th}$ percentile for all exposures) and 'healthy' (i.e., median value for all exposures). Red points denote risk reduction in increments of 10% (HR = 0.10) to the nearest value. Model is adjusted for age, sex, ethnicity, smoking, education, Townsend deprivation index, alcohol, discretionary screen time (time spent watching TV or using the computer outside of work), light intensity physical activity, medication (blood pressure, insulin, and cholesterol), previous diagnosis of major CVD (defined as disease of the circulatory system, arteries, and lymph, excluding hypertension), previous diagnosis of cancer, and familial history of CVD and cancer.

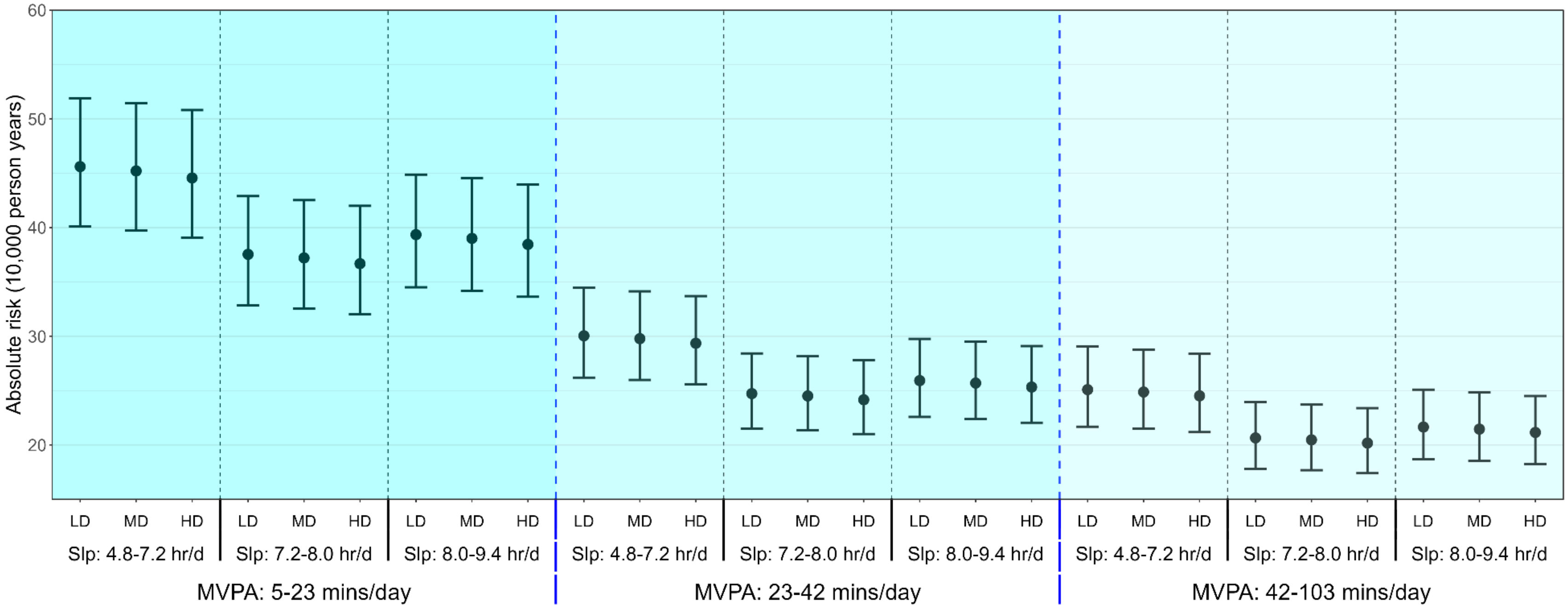


**Supplementary Figure 3:** Association of sleep, MVPA, and diet quality with absolute all-cause mortality risk

**Legend:** Model is adjusted for age, sex, ethnicity, smoking, education, Townsend deprivation index, alcohol, discretionary screen time (time spent watching TV or using the computer outside of work), light intensity physical activity, medication (blood pressure, insulin, and cholesterol), previous diagnosis of major CVD (defined as disease of the circulatory system, arteries, and lymph, excluding hypertension), previous diagnosis of cancer, and familial history of CVD and cancer. Dashed blue lines separate tertiles MVPA and dashed black lines separate tertiles of sleep. Moderate-Vigorous Physical Activity (MVPA); Low Diet Quality (LD); Medium Diet Quality (MD); High Diet Quality (HD).

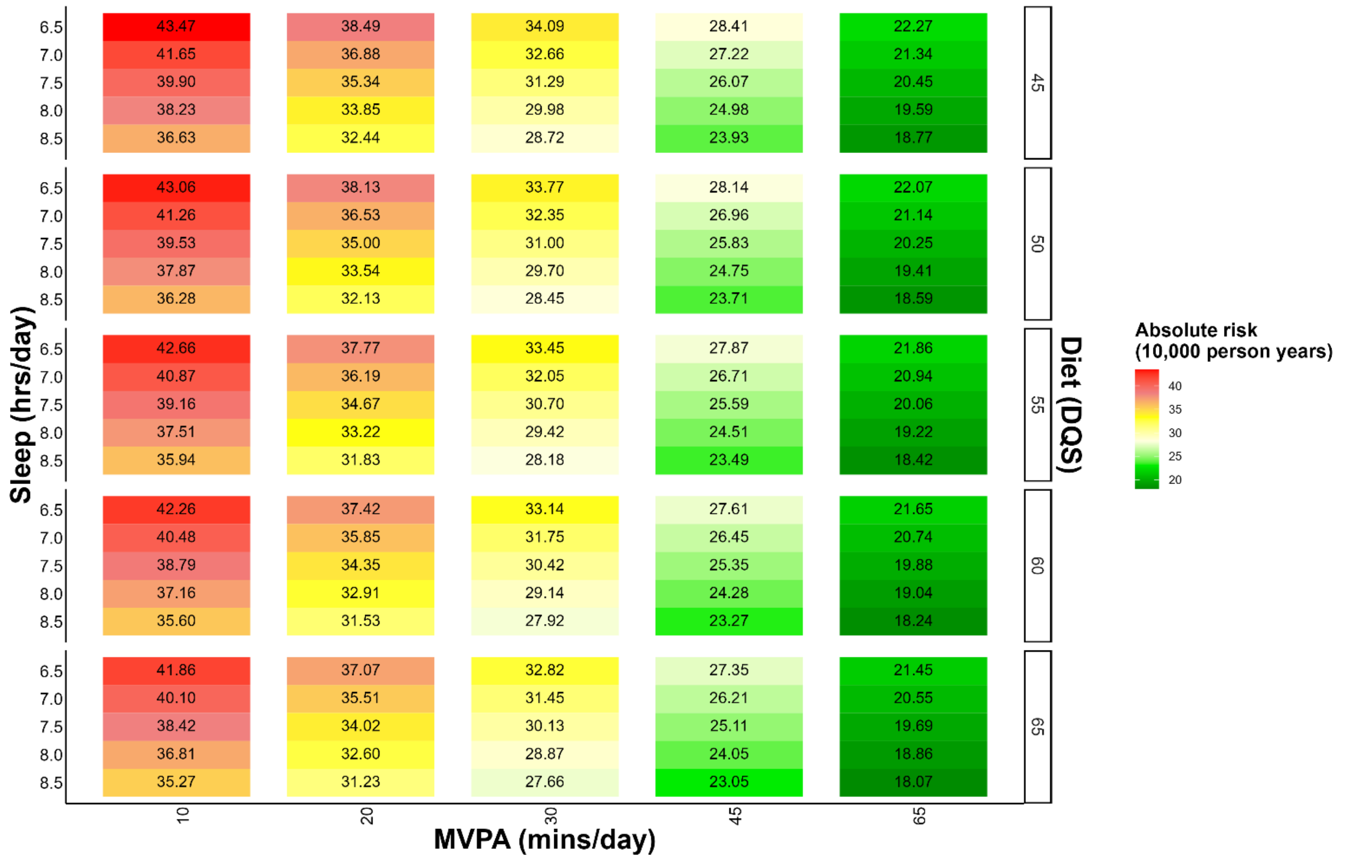


**Supplemental Figure 4:** Absolute all-cause mortality risk associated with concurrent variations in sleep, MVPA, and dietary quality score

**Legend**: The correlogram displays incremental changes in SPAN and absolute all-cause mortality risk as incidence per 10,000 person years with the reference being the 5th percentile of sleep (5.5 hours/day), physical activity (7.3 mins/day), and nutrition (36.9 DQS). Moderate-Vigorous Physical Activity (MVPA); Diet Quality Score (DQS). Model is adjusted for age, sex, ethnicity, smoking, education, Townsend deprivation index, alcohol, discretionary screen time (time spent watching TV or using the computer outside of work), light intensity physical activity, medication (blood pressure, insulin, and cholesterol), previous diagnosis of major CVD (defined as disease of the circulatory system, arteries, and lymph, excluding hypertension), previous diagnosis of cancer, and familial history of CVD and cancer.

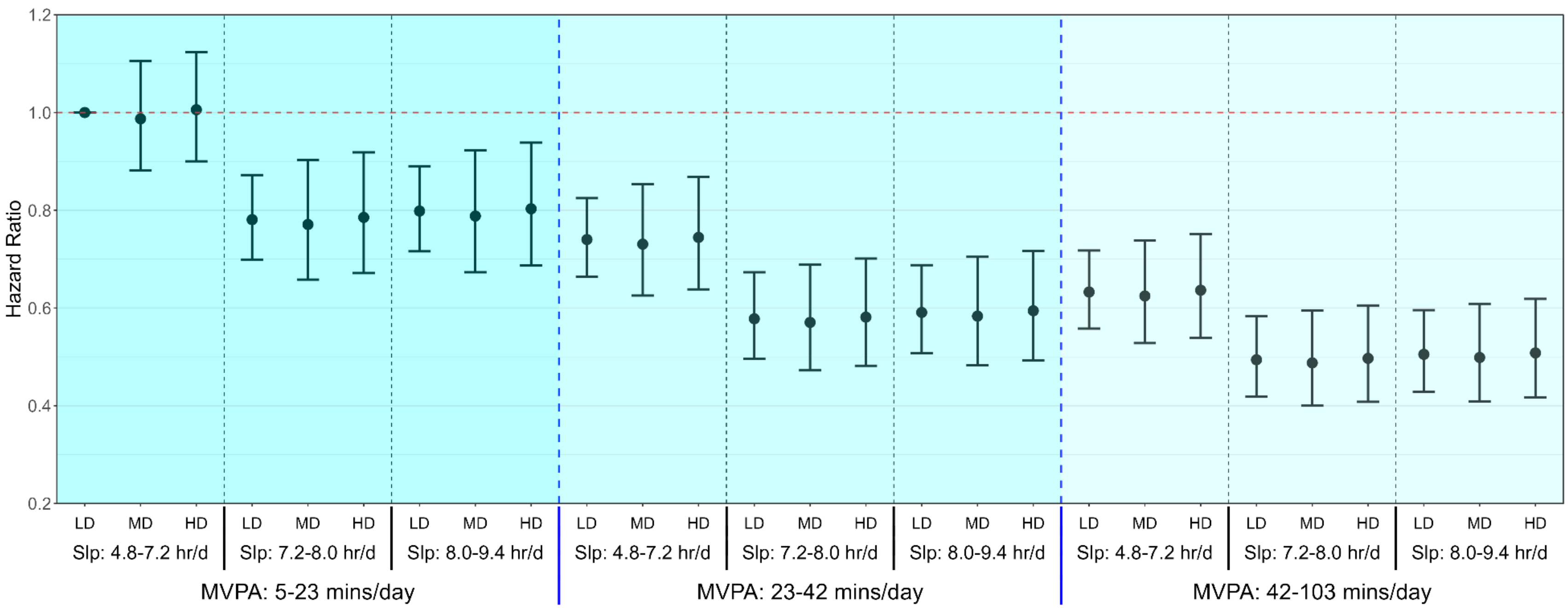


**Supplemental Figure 5:** Association of sleep, MVPA, and diet quality with all-cause mortality with exclusion of poor health individuals

**Legend**: Forest plot shows the SPAN associations with all-cause mortality after removing those with poor health status including, low BMI (<18.5), current smokers, self-reported poor health, and those with a frailty index score of >3. Model is adjusted for age, sex, ethnicity, smoking, education, Townsend deprivation index, alcohol, discretionary screen time (time spent watching TV or using the computer outside of work), light intensity physical activity, medication (blood pressure, insulin, and cholesterol), previous diagnosis of major CVD (defined as disease of the circulatory system, arteries, and lymph, excluding hypertension), previous diagnosis of cancer, and familial history of CVD and cancer. Dashed blue lines separate tertiles MVPA and dashed black lines separate tertiles of sleep. Moderate-Vigorous Physical Activity (MVPA); Low Diet Quality (LD); Medium Diet Quality (MD); High Diet Quality (HD).

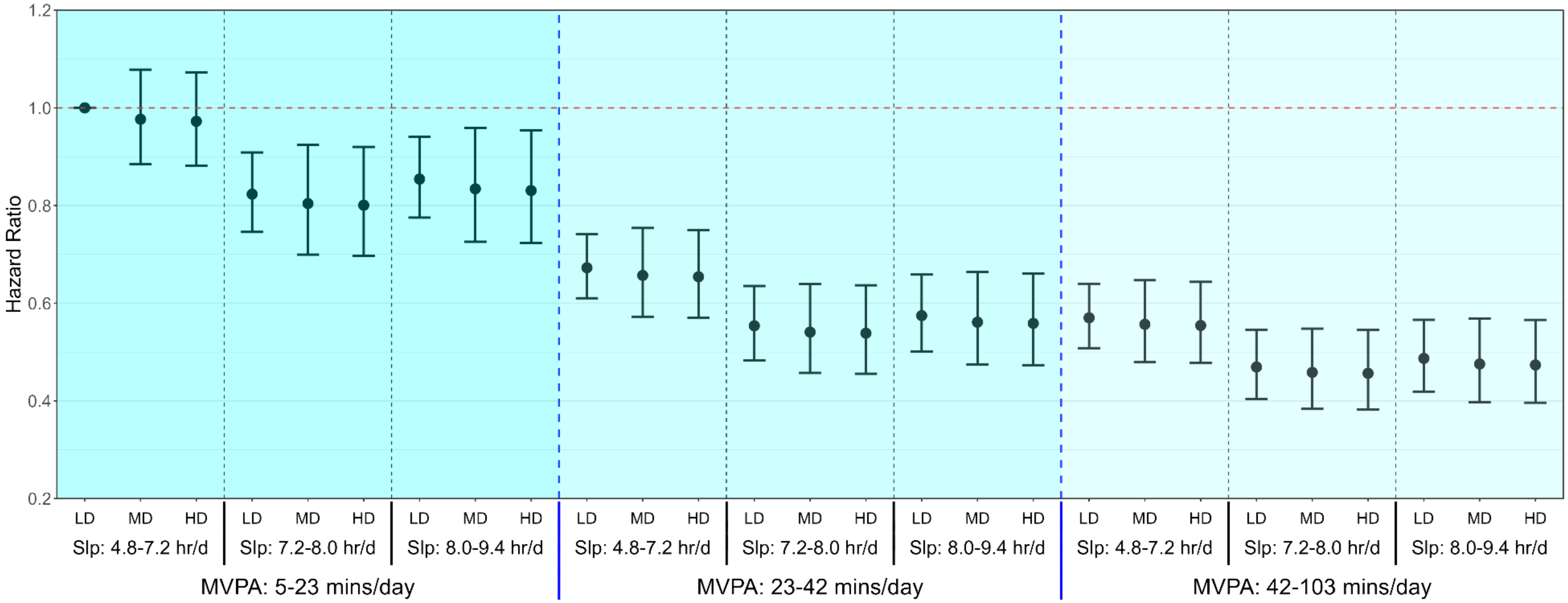


**Supplemental Figure 6:** Association of sleep, MVPA, and diet quality with all-cause mortality with adjustment for BMI
**Legend**: Model is adjusted for age, sex, ethnicity, smoking, education, Townsend deprivation index, alcohol, discretionary screen time (time spent watching TV or using the computer outside of work), light intensity physical activity, medication (blood pressure, insulin, and cholesterol), previous diagnosis of major CVD (defined as disease of the circulatory system, arteries, and lymph, excluding hypertension), previous diagnosis of cancer, familial history of CVD and cancer, and BMI. Dashed blue lines separate tertiles MVPA and dashed black lines separate tertiles of sleep. Moderate-Vigorous Physical Activity (MVPA); Low Diet Quality (LD); Medium Diet Quality (MD); High Diet Quality (HD).

**Supplementary Table 1:** Covariate Definitions.

| Variable | Definition | UK Biobank field ID (if applicable) |
|---|---|---|
| Age | Categorical (4) | 34, 52, accelerometer date-timestamp |
| Sex | Female/Male | 31 |
| Ethnicity | White/Others | 21000 |
| Education | College/University; A/AS level; O levels; CSE; NVQ/HND/HNC; other | 6138 |
| Smoking status | Never, past, current | 20116 |
| Alcohol consumption | Units/day | 20403 |
| Light intensity physical activity | Standing utilitarian movements, slow walking (<3 METs) | Derived from accelerometer data |
| Discretionary screen-time | Time spent/day watching TV and using a computer outside of work | 1070, 1080 |
| Townsend deprivation | Categorical (5) | 22189 |
| Use of cholesterol medication | Yes/No | 6177, 6153 |
| Use of blood pressure medication | Yes/No | 6177, 6153 |
| Use of diabetes medication | Yes/No | 6177, 6153 |
| Previous CVD | Identified by self-report and cancer registry. Defined as disease of the circulatory system, arteries, and lymph, excluding hypertension | 2001,100092 |
| Previous cancer | Identified by self-report and hospitalisation | 20002, 2000 |
| Familial history of CVD | Self-reporter mother of father diagnosed with heart disease or stroke | 20107, 20110 |
| Familial history of cancer | Self-reporter mother of father diagnosed with cancer | 20107, 20110 |
| High frailty scale | Categorical (yes/no); high frailty indicates a score of ≥3 on a 0 to 5 | 2306, 120107, 2624, 1011, 3637, 991, 971, 924, 46, 47 |
| Body mass index | Continuous; kilogram/meter$^2$ | Collected by trained professional |

**Supplementary Table 2:** STROBE Statement.

| | Item No | Recommendation | Page No |
|---|---|---|---|
| **Title and abstract** | 1 | (*a*) Indicate the study's design with a commonly used term in the title or the abstract | 1 |
| | | (*b*) Provide in the abstract an informative and balanced summary of what was done and what was found | 2 |
| **Introduction** | | | |
| Background/rationale | 2 | Explain the scientific background and rationale for the investigation being reported | 3 |
| Objectives | 3 | State specific objectives, including any prespecified hypotheses | 4-5 |
| **Methods** | | | |
| Study design | 4 | Present key elements of study design early in the paper | |
| Setting | 5 | Describe the setting, locations, and relevant dates, including periods of recruitment, exposure, follow-up, and data collection | 5 |
| Participants | 6 | (*a*) Give the eligibility criteria, and the sources and methods of selection of participants. Describe methods of follow-up | 5 |
| | | (*b*) For matched studies, give matching criteria and number of exposed and unexposed | - |
| Variables | 7 | Clearly define all outcomes, exposures, predictors, potential confounders, and effect modifiers. Give diagnostic criteria, if applicable | 6 |
| Data sources/ measurement | 8* | For each variable of interest, give sources of data and details of methods of assessment (measurement). Describe comparability of assessment methods if there is more than one group | 5 |
| Bias | 9 | Describe any efforts to address potential sources of bias | 7-8 |
| Study size | 10 | Explain how the study size was arrived at | 5, 8 |

| | | | |
|---|---|---|---|
| Quantitative variables | 11 | Explain how quantitative variables were handled in the analyses. If applicable, describe which groupings were chosen and why | 6 |
| Statistical methods | 12 | (*a*) Describe all statistical methods, including those used to control for confounding | 6-8 |
| | | (*b*) Describe any methods used to examine subgroups and interactions | 8 |
| | | (*c*) Explain how missing data were addressed | 5 |
| | | (*d*) If applicable, explain how loss to follow-up was addressed | 5 |
| | | (*e*) Describe any sensitivity analyses | 8 |
| **Results** | | | |
| Participants | 13* | (a) Report numbers of individuals at each stage of study—eg numbers potentially eligible, examined for eligibility, confirmed eligible, included in the study, completing follow-up, and analysed | 5 |
| | | (b) Give reasons for non-participation at each stage | Supplemental figure 1 |
| | | (c) Consider use of a flow diagram | Supplemental figure 1 |
| Descriptive data | 14* | (a) Give characteristics of study participants (eg demographic, clinical, social) and information on exposures and potential confounders | Table 1 |
| | | (b) Indicate number of participants with missing data for each variable of interest | Supplemental figure 1 |
| | | (c) Summarise follow-up time (eg, average and total amount) | Table 1 |
| Outcome data | 15* | Report numbers of outcome events or summary measures over time | 8 |

| | | | |
|---|---|---|---|
| Main results | 16 | (*a*) Give unadjusted estimates and, if applicable, confounder-adjusted estimates and their precision (eg, 95% confidence interval). Make clear which confounders were adjusted for and why they were included | 8-9 |
| | | (*b*) Report category boundaries when continuous variables were categorized | 8-9 |
| | | (*c*) If relevant, consider translating estimates of relative risk into absolute risk for a meaningful time period | 10 |
| Other analyses | 17 | Report other analyses done—eg analyses of subgroups and interactions, and sensitivity analyses | 11 |
| **Discussion** | | | |
| Key results | 18 | Summarise key results with reference to study objectives | 12 |
| Limitations | 19 | Discuss limitations of the study, taking into account sources of potential bias or imprecision. Discuss both direction and magnitude of any potential bias | 14-15 |
| Interpretation | 20 | Give a cautious overall interpretation of results considering objectives, limitations, multiplicity of analyses, results from similar studies, and other relevant evidence | 15 |
| Generalisability | 21 | Discuss the generalisability (external validity) of the study results | 15 |
| **Other information** | | | |
| Funding | 22 | Give the source of funding and the role of the funders for the present study and, if applicable, for the original study on which the present article is based | 16 |

*Give information separately for exposed and unexposed groups.

**Note:** An Explanation and Elaboration article discusses each checklist item and gives methodological background and published examples of transparent reporting. The STROBE checklist is best used in conjunction with this article (freely available on the Web sites of PLoS Medicine at http://www.plosmedicine.org/, Annals of Internal Medicine at http://www.annals.org/, and Epidemiology at http://www.epidem.com/). Information on the STROBE Initiative is available at http://www.strobe-statement.org.
